# The impact of polygenic score and socioeconomic status in predicting risk for 19 complex diseases

**DOI:** 10.1101/2025.09.02.25334913

**Authors:** Fiona A. Hagenbeek, Anne Richmond, Max Tamlander, Kira Detrois, Zhiyu Yang, Tuomo Hartonen, Daniel L. McCartney, FinnGen, Riccardo E. Marioni, Pekka Martikainen, Nina Mars, Andrea Ganna, Samuli Ripatti

## Abstract

Both socioeconomic circumstances and genetic predisposition shape disease risk, yet their joint contribution across diseases has not been systematically examined. We studied 19 high-burden diseases in 743,194 participants (729,928 European; 13,266 non-European ancestry) from FinnGen, the UK Biobank, and Generation Scotland. Higher educational attainment was associated with lower risk of most conditions, but with higher risk of most common cancers. These associations were largely independent of disease-specific polygenic scores (PGSs). For seven out of 19 diseases, PGSs showed stronger effects among individuals with high education. Joint inclusion of education and PGSs modestly improved prediction for 14 and 10 out of 19 diseases in FinnGen and the UK Biobank, respectively. PGS associations were consistent across ancestries, whereas education effects were less stable; results using an alternative socioeconomic measure were directionally similar but smaller. Our findings highlight the distinct and partly interacting contributions of socioeconomic and genetic factors to disease risk.

---

Socioeconomic inequalities are linked to increased disease burden for many complex diseases^1–3^. Most diseases have higher incidence among individuals with low socioeconomic status (SES), such as cardiometabolic and mental health disorders^1,2^. For example, individuals without formal schooling have a 30% higher overall disease burden than individuals with higher educational attainment^3^. In contrast, cancers, particularly breast cancer and skin melanoma, are more prevalent in individuals with high SES^2,4,5^, especially in older age groups^1^.

In addition to socioeconomic differences, genetic factors affect complex disease susceptibility, and genome-wide association studies (GWAS) have successfully identified thousands of common variant associations for complex traits and diseases^6^. The availability of large GWAS results have led to the development of polygenic scores (PGS), which sum the effects of many common genetic variants of small effect sizes into a single score predictive of inherited disease risk or complex trait outcomes^7,8^. PGSs significantly predict diseases^9,10^, and have been shown to provide additional information for disease risk when considered alongside clinical risk factors^11,12^ or family history^13^. Also, socioeconomic measures have substantial environmental influences but also genetic influences, particularly for educational attainment^14^. PGSs for educational attainment predict common diseases such as major depression, type 2 diabetes (T2D), and heart disease, and including the PGS for educational attainment, the disease-specific PGS, and their interaction substantially increases disease prediction^14^. These results show that genetic factors are associated with both socioeconomic factors and complex disease risks.

While PGSs capture inherited susceptibility, genetic risk does not imply determinism^15^. Evidence from coronary artery disease and T2D shows that lifestyle interventions can meaningfully reduce risk in genetically high-risk individuals^16,17^. These findings underscore the value of integrating genetic and environmental risk information to inform preventive strategies. In the current study, we aim to explore the associations between PGSs and SES, as measured by educational attainment, on complex disease risk for 19 high-burden diseases^18^ with known socioeconomic and genetic differences in risk across three studies in Finland and the United Kingdom (**N** = 729,928). We set to test the following hypotheses of public health importance: 1) both PGSs and SES are independently associated with 19 common diseases, 2) the strength of the association between PGSs and diseases differs across levels of SES, and 3) models that include both PGSs and SES provide better predictive ability for complex diseases than models including only one of these factors.

## Results

### Association of PGSs, education, and their interaction with 19 diseases

We harmonized disease and education definitions across FinnGen (*N* = 360,663), UK biobank (*N* = 353,294 European and 13,266 non-European ancestry), and Generation Scotland (*N* = 15,971), and calculated PGSs using a previously reported pipeline^19^ (see **Methods**). Unless otherwise specified, all results are provided for participants of European ancestry. Descriptive statistics for participants of European ancestry are shown in **Table 1** (**Supplementary Table 4**).

**Table 1.** Descriptive statistics for participants of European ancestry by study and complex disease. Follow-up time is defined as age from birth until the end of the last linking between the registry and biobank study. Descriptive statistics by case-control status and educational attainment for each complex ease in each biobank study are in Supplementary Table 4.

| Study | Disease | Sample size (N) | Age of recruitment (years) mean (IQR) | Maximum follow-up time across diseases (years) median (IQR) | Disease prevalence | N (%) females | N (%) high education | Ascertainment strategy |
| --- | --- | --- | --- | --- | --- | --- | --- | --- |
| FinnGen | Type 1 Diabetes | 281691 | 48 (18.6) | 60.64 (20.42) | 0.3% | 164440 (58.4%) | 130164 (46.2%) | Population and hospital |
|  | Prostate Cancer | 156812 | 68.1 (10.1) | 65.36 (18.18) | 9.9% | 0 (0%) | 56547 (36.1%) |  |
|  | Type 2 Diabetes | 339250 | 60.4 (14.6) | 63.15 (19.97) | 18.8% | 188656 (55.6%) | 145348 (42.8%) |  |
|  | Gout | 324123 | 64.8 (16.3) | 62.16 (20) | 2.8% | 178382 (55%) | 141921 (43.8%) |  |
|  | Rheumatoid Arthritis | 345393 | 56.7 (17.2) | 62.37 (19.81) | 2.7% | 192829 (55.8%) | 149647 (43.3%) |  |
|  | Breast Cancer | 195041 | 58.8 (15.5) | 60.38 (20.94) | 9.9% | 195041 (100%) | 95333 (48.9%) |  |
|  | Atrial Fibrillation | 226332 | 67.3 (13.9) | 59.22 (23) | 20.1% | 129285 (57.1%) | 106463 (47%) |  |
|  | Colorectal Cancer | 310692 | 66.8 (13.6) | 61.75 (20.1) | 2.1% | 174701 (56.2%) | 137121 (44.1%) |  |
|  | Asthma | 206120 | 56.3 (19) | 63.21 (19.42) | 14.9% | 119964 (58.2%) | 92504 (44.9%) |  |
|  | Coronary Heart Disease | 332614 | 64.5 (14.9) | 62.71 (20.5) | 12.4% | 183558 (55.2%) | 143117 (43%) |  |
|  | Hip Osteoarthritis | 285508 | 64.3 (14.4) | 61.43 (20.99) | 8.7% | 156590 (54.8%) | 127683 (44.7%) |  |
|  | Knee Osteoarthritis | 311651 | 59.1 (15.3) | 62.45 (20.44) | 16.4% | 173468 (55.7%) | 135266 (43.4%) |  |
|  | Skin Melanoma | 345238 | 62 (17.8) | 62.28 (19.81) | 1.0% | 192699 (55.8%) | 150021 (43.5%) |  |
|  | Lung Cancer | 238688 | 70.5 (9.5) | 61.38 (20.59) | 2.0% | 133623 (56%) | 105010(44%) |  |
|  | Major Depression | 331350 | 50.3 (14.5) | 62.99 (19.01) | 9.8% | 183505 (55.4%) | 144774 (43.7%) |  |
|  | Any Cancer | 360663 | 64.4 (15.1) | 63.37 (20.29) | 24.4% | 198569 (55.1%) | 153728 (42.6%) |  |
|  | Epilepsy | 263783 | 58.3 (21.2) | 61.96 (20.46) | 2.9% | 148000 (56.1%) | 116970 (44.3%) |  |
|  | Appendicitis | 327734 | 48.9 (17.4) | 62.81 (19.75) | 4.6% | 182216 (55.6%) | 141519 (43.2%) |  |
|  | Alcohol Use Disorder | 305220 | 51.9 (17.8) | 61.96 (19.92) | 5.2% | 172354 (56.5%) | 136142 (44.6%) |  |
| UK Biobank | Type 1 Diabetes | 352675 | 55.8 (15.4) | 68.45 (12.5) | 0.2% | 192462 (54.6%) | 139031 (39.4%) | Population |
|  | Prostate Cancer | 161706 | 67.4 (9.4) | 68.96 (12.67) | 8.0% | 0 (0%) | 66232 (41%) |  |
|  | Gout | 353294 | 68.5 (11.8) | 68.54 (12.51) | 2.0% | 192588 (54.5%) | 139217 (39.4%) |  |
|  | Rheumatoid Arthritis | 352733 | 64 (13.2) | 68.45 (12.5) | 0.2% | 192498 (54.6%) | 139051 (39.4%) |  |
|  | Breast Cancer | 193453 | 59 (14.5) | 68.29 (12.42) | 8.1% | 193453 (100%) | 73748 (38.1%) |  |
|  | Epilepsy | 352862 | 61.2 (17) | 68.54 (12.5) | 1.3% | 192531 (54.6%) | 139095 (39.4%) |  |
|  | Alcohol Use Disorder | 352882 | 61.5 (16) | 68.54 (12.5) | 2.0% | 192511 (54.6%) | 139121 (39.4%) |  |
| Generation Scotland | Type 1 Diabetes | 15765 | 49.3 (15.2) | 61.24 (19.22) | 0.3% | 9261 (58.7%) | 5378 (34.1%) | Population |
|  | Prostate Cancer | 6532 | 66.9 (8.2) | 61.61 (20.04) | 3.1% | 0 (0%) | 2177 (33.3%) |  |
|  | Type 2 Diabetes | 15826 | 64 (13.6) | 61.28 (19.24) | 3.2% | 9285 (58.7%) | 5384 (34%) |  |
|  | Breast Cancer | 9295 | 57 (13.3) | 61.07 (18.69) | 4.8% | 9295 (100%) | 3212 (34.6%) |  |
|  | Asthma | 15598 | 57.1 (18.2) | 61.5 (19.04) | 3.6% | 9171 (58.8%) | 5314 (34.1%) |  |
|  | Coronary Heart Disease | 15971 | 60.6 (14.6) | 61.45 (19.41) | 6.8% | 9356 (58.6%) | 5406 (33.8%) |  |
|  | Hip Osteoarthritis | 15841 | 64.1 (11.7) | 61.28 (19.26) | 2.9% | 9308 (58.8%) | 5388 (34%) |  |
|  | Knee Osteoarthritis | 15856 | 64.4 (12.2) | 61.31 (19.28) | 2.9% | 9323 (58.8%) | 5387 (34%) |  |
|  | Skin Melanoma | 15780 | 61.9 (19.6) | 61.23 (19.23) | 0.4% | 9271 (58.8%) | 5379 (34.1%) |  |
|  | Lung Cancer | 15785 | 65.7 (9.6) | 61.22 (19.24) | 0.8% | 9276 (58.8%) | 5379 (34.1%) |  |
|  | Major Depression | 15735 | 52.1 (15.1) | 61.27 (19.19) | 1.1% | 9238 (58.7%) | 5373 (34.1%) |  |
|  | Any Cancer | 15901 | 61.5 (15.1) | 61.45 (19.39) | 10.7% | 9328 (58.7%) | 5395 (33.9%) |  |
|  | Epilepsy | 15728 | 55.9 (14.3) | 61.27 (19.19) | 0.7% | 9240 (58.7%) | 5366 (34.1%) |  |
|  | Appendicitis | 15517 | 48.9 (18.5) | 61.44 (19.13) | 1.0% | 9125 (58.8%) | 5296 (34.1%) |  |
|  | Alcohol Use Disorder | 15717 | 53.3 (15.3) | 61.28 (19.19) | 1.3% | 9235 (58.8%) | 5371 (34.2%) |  |

IQR, interquartile range

We evaluated the association between PGSs, education, and their interaction with 19 diseases with Cox proportional hazard models in each study followed by fixed-effect meta-analysis across the studies. All PGSs were significantly associated with their respective diseases (**Fig. 1A**, **Supplementary Table 5**), with hazard ratios (HR) per 1 standard deviation (SD) of PGS ranging from 2.11 (95% confidence interval (CI): 2.01-2.21) for type 1 diabetes (T1D) to 1.06 (95% CI: 1.04-1.07) for alcohol use disorder (AUD). Associations were largely consistent across cohorts, though we observed heterogeneity for several diseases, including knee osteoarthritis (*Q* = 11.97, *p* = 5.40×10^-04^), T2D (*Q* = 13.61, *p* = 2.25×10^-04^), prostate cancer (*Q* = 16.67, *p* = 2.41×10^-04^), gout (*Q* = 44.78, *p* = 2.21×10^-11^), asthma (*Q* = 45.28, *p* = 1.70×10^-11^), and rheumatoid arthritis (RA, *Q* = 60.43, *p* = 7.62×10^-15^), based on Cochran’s *Q* test (Bonferroni-adjusted *p* < 2.63×10^-03^, **Supplementary Table 5**, **Supplementary Fig. 3**). Associations were not attenuated after adjustment for high education (**Supplementary Tables 5-7**, **Supplementary Fig.s 3**, **4A**, and **5**).

**Fig. 1.**
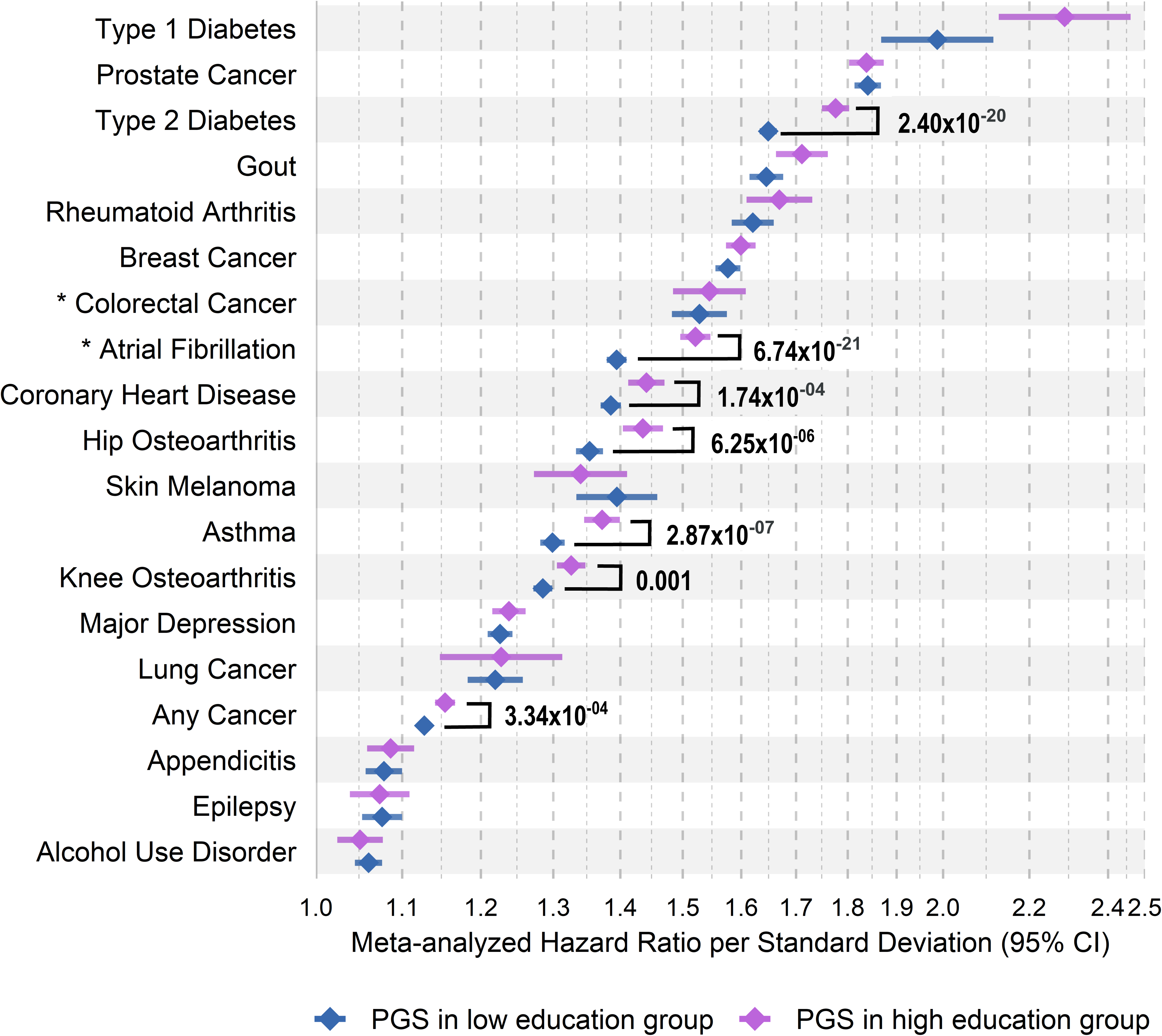
Meta-analyzed hazard ratios for the relative risk of educational attainment and disease-specific polygenic scores on 19 complex diseases. A. Mixed-effects meta-analyzed hazard ratios per standard deviation of the disease-specific polygenic scores on risk of complex diseases. B. Mixed-effects meta-analyzed hazard ratios for individuals with high education relative to individuals with low education on risk of complex diseases. The asterisk indicates the complex disease was not meta-analyzed and assessed only in the FinnGen study. The exact values are in **Supplementary Table 5**. Sample sizes by case-control status and educational attainment for each complex disease in each biobank study are in **Supplementary Table 4**.

High education was associated with lower risk for 12 diseases, including T2D, coronary heart disease (CHD), asthma, and osteoarthritis (HR range, 0.58–0.85 [95% CI range, 0.54-0.88]), and with increased risk for skin melanoma, breast, colorectal, prostate, and any cancer (HR range, 1.10–1.40 [95% CI range, 1.04-1.50]), and was not associated with risks of T1D or appendicitis (**Fig. 1A**, **Supplementary Table 5**). These associations also showed heterogeneity for T2D (*Q* = 13.44, *p* = 2.46×10^-04^), prostate cancer (*Q* = 15.45, *p* = 4.41×10^-04^), gout (*Q* = 19.96, *p* = 7.91×10^-06^), AUD (*Q* = 55.59, *p* = 8.49×10^-13^), and breast cancer (*Q* = 80.16, *p* = 3.93×10^-18^, **Supplementary Table 5**, **Supplementary Fig. 3**). Adjusting for PGSs had minimal impact on education effects, except for T2D (HR difference = 0.03, *p* = 0.0001; **Fig. 1B**, **Supplementary Tables 5-7**, **Supplementary Fig.s 3**, **4B**, and **5**), suggesting independent contributions of genetic risk and education to disease risk.

We next estimated the association of PGSs by stratifying on education level and found that for seven diseases, including atrial fibrillation (AF), T2D, CHD, osteoarthritis, asthma, and any cancer, PGS effect sizes were larger among individuals with high education (Bonferroni-adjusted *p* < 2.63×10^-03^; **Fig. 2**, **Supplementary Table 8-9**). For example, for AF, the PGS HRs per SD were 1.39 (95% CI: 1.38–1.41) among individuals with low education and 1.52 (95% CI: 1.50–1.55) among those with high education. For T2D, HRs were 1.65 (95% CI: 1.63–1.66) and 1.78 (95% CI: 1.75–1.80), respectively. Heterogeneity across cohorts in these stratified analyses was evident in the low education group for asthma (*Q* = 26.65, *p* = 2.43×10^-07^), prostate cancer (*Q* = 26.88, *p* = 1.46×10^-06^), gout (*Q* = 32.11, *p* = 1.45×10^-08^), and RA (*Q* = 46.08, *p* = 1.13×10^-11^), and in the high education group for gout (*Q* = 9.92, *p* = 1.64×10^-03^), appendicitis (*Q* = 10.75, *p* = 1.04×10^-03^), RA (*Q* = 13.98, *p* = 1.85×10^-04^), and asthma (*Q* = 21.87, *p* = 2.91×10^-06^, **Supplementary Table 8**, **Supplementary Fig. 6**).

**Fig. 2.**
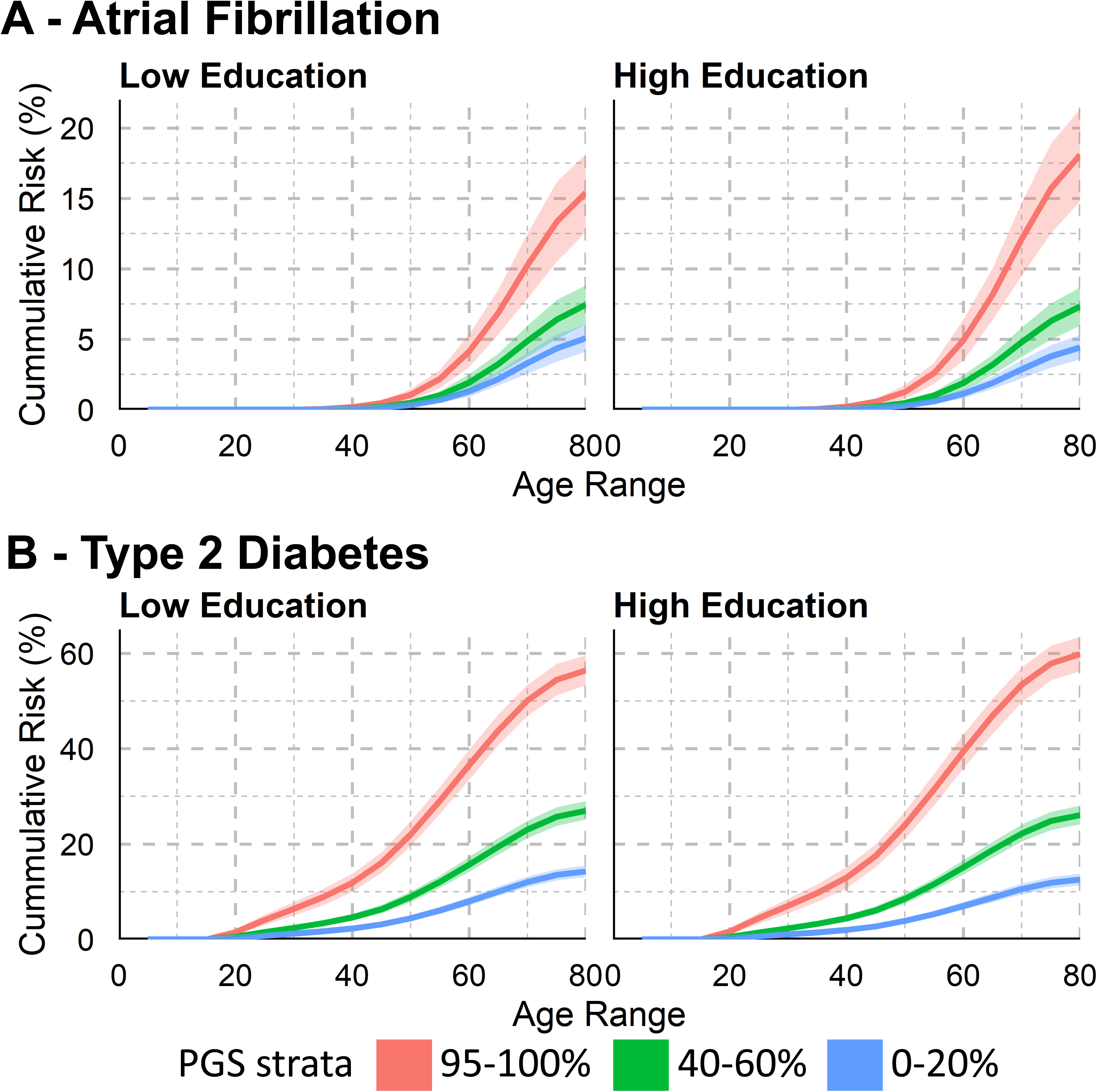
Meta-analyzed hazard ratios for the relative risk of the disease-specific polygenic scores by educational attainment on 19 complex diseases. Mixed-effects meta-analysis of the log hazard ratios per standard deviation of the disease-specific polygenic score in the low (blue) or high (purple) education group. The asterisk indicates the complex disease was not meta-analyzed and assessed only in the FinnGen study. Significance of the differences in the effect of the disease-specific polygenic score per education group evaluated by the statistical significance of the interaction term between the disease-specific PGS and education level after Bonferroni correction for multiple testing of 19 outcomes (*p* < 2.63×10^-03^). The exact values are in **Supplementary Tables 8-9**. Sample sizes by case-control status and educational attainment for each complex disease in each biobank are in **Supplementary Table 4**.

### Cumulative incidence estimation stratified by PGS and education level

For each disease for which we observed a significant interaction between PGSs and education in FinnGen, we derived Finnish-specific estimates of the cumulative incidence by PGS quantiles, stratified by education level, where the baseline risk was calibrated using the Global Burden of Disease (GBD)^18^. Cumulative incidence curves demonstrated that disease risk increased with higher PGS levels and differed by education (**Fig. 3, Supplementary Tables 10-12**, **Supplementary Fig. 7**). For instance, individuals in the top 5% of the PGS distribution for AF or T2D with high education had higher cumulative incidence by age 80 compared to those with low education (**Fig. 3**, AF: 18.09% [95% CI: 14.79-21.36%] vs 15.41% [95% CI: 12.61-18.16%], T2D: 59.83% [95% CI: 56.31-63.47%] vs 56.44% [95% CI: 53.32-59.62%]).

**Fig. 3.**
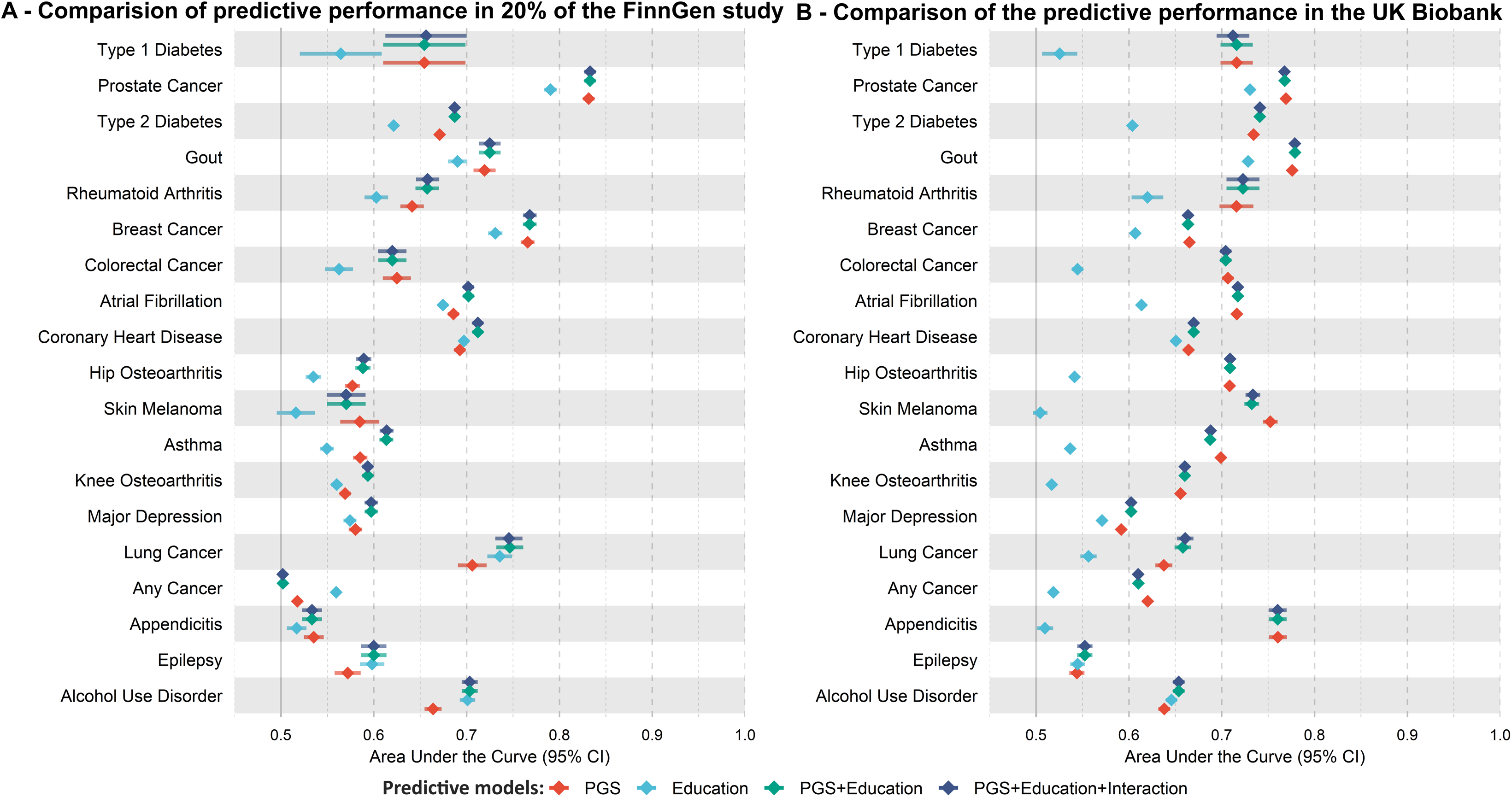
Educational attainment-specific cumulative incidence estimates in FinnGen. Bootstrapped 95% confidence intervals reflect the uncertainty of the cumulative incidence estimates for the top, median, and bottom of the disease-specific PGS distribution for (**A**) atrial fibrillation and (**B**) type 2 diabetes. The exact values are in **Supplementary Table 12**, the exact values of the underlying Cox proportional hazard model are in **Supplementary Table 11**, and the sample sizes by case-control status and educational attainment are in **Supplementary Table 10**.

### Effects of PGSs, education, and their interaction on disease prediction

To assess the added predictive value of combining genetic and education information, we trained logistic regression models in FinnGen and evaluated model performance in held-out data, with replication in the UK Biobank (see **Methods**). In FinnGen, models including both PGS and education outperformed models with education alone for all diseases except any cancer, while the combined model outperformed PGS alone models for 14 out the 19 diseases (**Figure 4**, **Supplementary Tables 13-19**). For example, in AF, education and PGS together had an Area Under the Curve (AUC) of 0.702 (95% CI: 0.696, 0.708), compared to 0.686 (95% CI: 0.680,0.692) for PGS alone and 0.675 (95% CI: 0.669,0.680) for education alone. Although absolute improvements in AUC were modest (e.g., +0.016 for PGS + education vs. PGS alone in AF), they were consistent across most diseases and confirmed by continuous net reclassification and integrated discrimination improvement metrics (**Supplementary Table 20**). We also observed improved prediction of the combined PGS and education model compared to education only in the UK Biobank for all studied diseases, though the combined model out performed PGS only models for fewer diseases in the UK Biobank (10 out of 19). Adding a PGS by education interaction term provided no predictive benefit across diseases in either FinnGen or the UK Biobank. For instance, in FinnGen the AUC for AF was 0.702 (95% CI 0.696– 0.708) with and without the interaction term. Together, these findings indicate that education contributes complementary, though incremental, predictive information beyond polygenetic risk for most complex diseases.

**Fig. 4.**
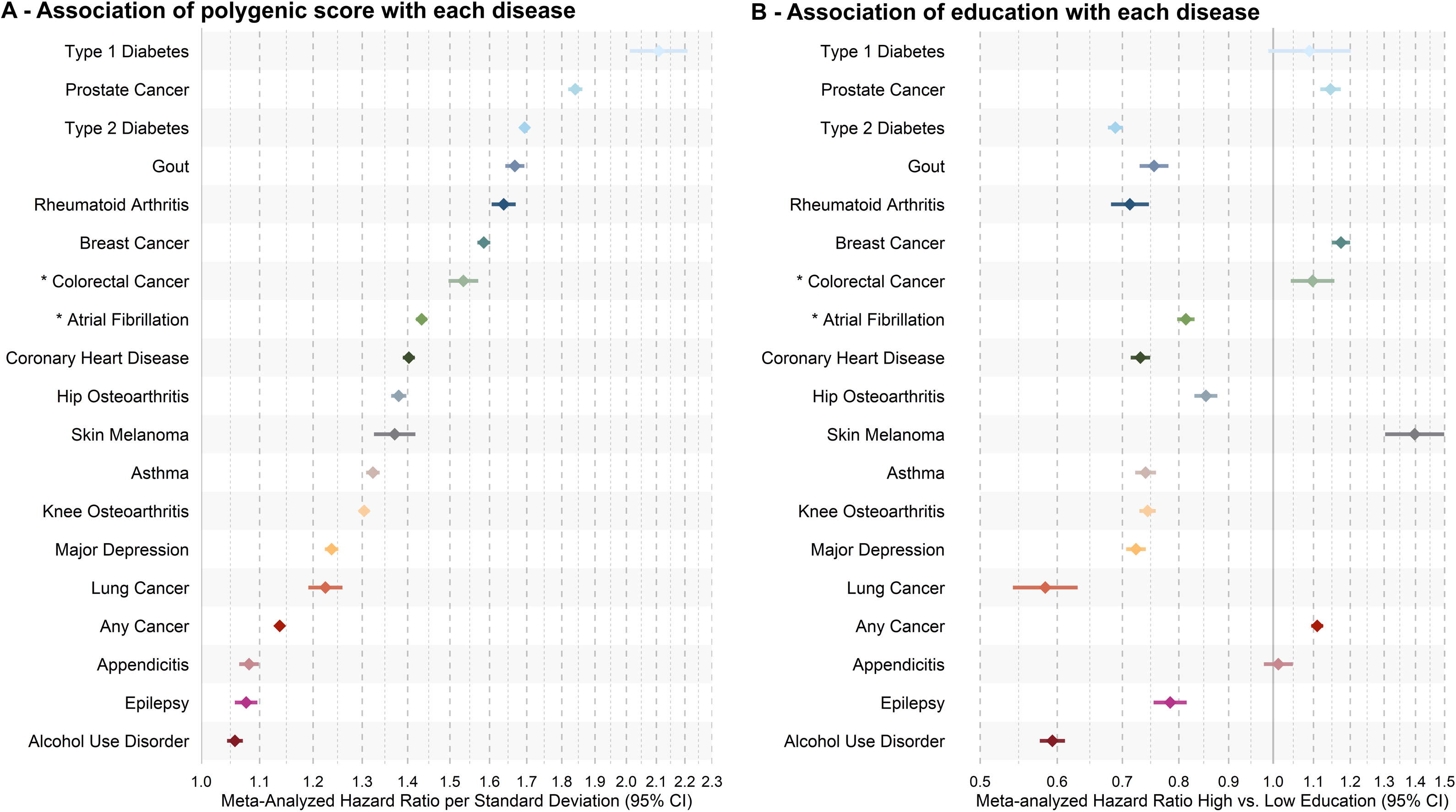
Predictive performance of disease-specific polygenic scores, educational attainment, and their combination. Area Under the Curve (AUC) with 95% confidence interval (95% CI) are show for predictive models in 20% of the FinnGen study (**A**) and the UK Biobank (**B**). Models include: 1) disease-specific polygenic score (PGS) only, 2) educational attainment (EA) only, 3) disease-specific PGS + EA, and 4) disease-specific PGS + EA + interaction disease-specific PGS and EA. AUCs were estimated from logistic regression model trained in 80% of the FinnGen study (see **Methods**, **Supplementary Tables 14-17**). Descriptive statistics by case-control status and educational attainment for each complex disease in 20% of the FinnGen study and all UK Biobank traits are in **Supplementary Table 18**. Full model estimates, including covariate-only baselines, are reported in **Supplementary Table 19**.

### Sensitivity analyses

Sensitivity analyses in FinnGen tested the robustness of the interaction findings by accounting for non–cause-specific mortality as a competing risk. The interaction effects remained unchanged, though the main effects of PGS and education were modestly attenuated (**Supplementary Fig. 8**, **Supplementary Tables 9, 21-23**). In the UK Biobank, we compared results between individuals of European and non-European ancestry (**Supplementary Methods**). PGS effects were generally consistent across ancestries, while education showed inconsistent associations in non-European ancestries, with wide CIs (**Supplementary Fig. 9, Supplementary Tables 5, 24-25**). Finally, we assessed occupational status as an alternative socioeconomic indicator in FinnGen (**Supplementary Methods**). Estimates were mostly consistent, but smaller in absolute size (**Supplementary Fig. 10-15, Supplementary Tables 26-34**), though only prostate cancer showed a significant difference in PGS effects by occupation (**Supplementary Fig. 11**, **Supplementary Tables 8-9** and **30-31**).

## Discussion

In this study, we assessed the association between SES, using educational attainment as a proxy, and PGSs for 19 complex diseases on the risk of these diseases and evaluated their predictive ability using data from 743,194 study participants across three biobank studies from Finland and the United Kingdom. We quantified the independent associations of both education and PGSs with the 19 common diseases. We showed that for CHD, T2D, AF, osteoarthritis, asthma, and any cancer the effect of disease-specific PGSs varies by education level. Additionally, combining PGSs and SES improved disease prediction compared to using either factor alone for most diseases, but their interaction did not enhance prediction models.

These findings allow us to draw several conclusions. First, SES, as measured by educational attainment, and PGSs both strongly affect the disease risks across many common diseases. The influence of SES on health outcomes has been extensively studied, and our findings are consistent with previous work demonstrating how lower SES contributes to susceptibility for premature mortality^20^ and chronic diseases like cardiovascular disease^21,22^. Conversely, higher SES has been associated with increased risk for many types of cancer, such as prostate cancer^23^, which may be partially explained by longer life expectancies for individuals with high SES^24^ which increases their exposure to age-related cancer risk^25^. However, the association between higher education and cancer risk may also be influenced by individuals with higher SES being more likely to undergo regular screening, resulting in earlier detection and higher observed incidence rates^26^.

Second, our findings show that both PGSs and education are independently associated with the risk of most diseases. Third, the influence of PGSs on disease risk varied by education level, consistent with previous findings for CHD^27,28^. This suggests that the expression of genetic risk depends on contextual factors, such as SES, lifestyle, and environmental exposures, rather than being uniformly expressed across populations. Specifically, in the context of our results, high educational attainment, often associated with more favorable social and lifestyle factors, may facilitate a stronger expression of genetic predispositions^29^. Finally, our study found that combining PGSs and education generally enhances disease prediction and highlight the importance of incorporating both genetic and social factors into risk prediction. However, it is important to remember that SES is a dynamic social construct, which is shaped by various factors, including genetically influenced traits, and impacts the distribution of genetic predispositions^29^. While SES contributes to identification of high-risk individuals, its causal role is unclear which needs to be considered when using SES in clinical settings.

There are many potential explanations for our observations. Individuals with lower SES tend to encounter more challenging environmental and social circumstances, potentially magnifying their genetic susceptibility to disease due to increased exposure to stress^30,31^, poorer access to healthcare, and limited health-promoting resources^32,33^. For example, SES-related differences in behaviors such as smoking and alcohol consumption contribute to many diseases including CHD^34^ and T2D^35^ and have been shown to modify the effects of genetic risk^16,17^. Conversely, individuals with higher SES are more likely to access health-promoting resources, adopt healthier lifestyles^32,33^, have greater health literacy^36,37^, and make more use of preventive healthcare^26,32,33^, which can mitigate genetic risk. In general, differences in SES, both at lower and higher levels, shape health behaviors and literacy, psychosocial stress, access to and likelihood of seeing medical services, and influence lifestyle factors that serve as intermediates in disease development, ultimately impacting the expression of genetic predispositions to diseases.

Our study has some limitations. First, our results were primarily based on European ancestry individuals living in Europe and need to be more extensively tested in non-European countries and mode diverse genetic ancestries to evaluate generalizability across diverse populations^38^. This is particularly relevant for countries with significant immigrant populations with non-European ancestry, as PGSs are less developed for non-Europeans^9^ but also as immigrants are often exposed to increased stress and adversities, regardless of their educational attainment^39^. However, our results in non-European ancestry individuals in the UKB were mostly in line with our observations in European ancestry individuals. Second, while we cover two different health care and educational systems, Finland and the UK are relatively equal in their health care access, with both countries having nationalized publicly available health care^40,41^, and universal educational access^42^. The dynamics between education and polygenic risk may be different in countries with varying degrees of healthcare coverage and educational accessibility^43,44^. Third, we primarily used educational attainment as our measure of SES as it is relatively simple to standardize across countries and educational systems^45^. However, we dichotomized education into high versus low attainment, which may oversimplify the spectrum of educational experiences and obscure more nuanced associations with polygenic risk. While we observed similar results when analyzing occupation, different socioeconomic indices, such as income or neighborhood derivation, may capture other facets of socioeconomic environments^46^, providing insight into the overlapping and unique aspects of the socioeconomic environment and how they might influence disease risk.

In summary, our findings indicate that both genetic and socioeconomic factors independently associate with disease risk, with context-dependent effect sizes. While integrating these factors into risk models could modestly improve predictive precision and inform targeted interventions, the observed improvements in our study were limited. Caution is warranted in extrapolating these results to claims about broader health equity, and future efforts should prioritize more diverse populations to ensure generalizability across different socioeconomic groups. Importantly, while socioeconomic indicators such as educational attainment are useful for population-level risk stratification, they should not be used in isolation to guide individual clinical decisions, as this could inadvertently worsen health inequities or contribute to stigmatization.

## Methods

### Participating studies

We analyzed data from 729,928 European ancestry participants from three studies across Finland and the United Kingdom (FinnGen^47^, UK Biobank^48^, and Generation Scotland^49^), as well as from 13,266 non-European ancestry participants from the UK Biobank. Participants were included if they were aged 35 to 80 at the time of study entry, to ensure most individuals had completed their formal education. Details on genotyping, imputation, quality control, and ancestry assignment for each cohort are available in the **Supplementary Methods**.

### Selection and harmonization complex diseases and educational attainment

We examined 19 diseases, selected based on their high burden in high-income countries according to the Global Burden of Disease^18^ and the availability of well-powered GWAS. Eighteen of these diseases were previously analyzed by the INTERVENE consortium^19^. We additionally included alcohol use disorder, given its known socioeconomic disparities in outcomes and high public health burden^50^, particularly in Finland^18^. Moreover, large-scale GWAS summary statistics for alcohol consumption are available^51^, allowing comparable PGS estimation.

Disease case definitions followed Jermy et al. (2024)^19^: participants were classified as cases based on the presence of harmonized ICD-9 or ICD-10 codes, as curated by clinical experts in FinnGen^47^. Controls were defined as individuals without those codes. Diagnoses were extracted from the health registries or electronic health records and were thus not limited to only information from after study enrollment (see **Supplementary Methods**).

To harmonize education across countries, we converted national education levels to the 1997 International Standard Classification of Education (ISCED)^45^. We then dichotomized education into low (ISCED ≤ 4) and high (ISCED ≥ 5) categories (see **Supplementary Table 2**). This cutoff was based on the distribution of education in FinnGen, chosen to ensure sufficient sample sizes and adequate case numbers within each group for all outcomes.

### Estimating polygenic scores

PGSs for 18 diseases were sourced from the INTERVENE consortium^19^ and are available via the PGS Catalog^52^ (publication ID PGP000618; score IDs PGS004869–PGS004886, **Supplementary Table 3**). For AUD, we used the same approach as INTERVENE to create a new PGS based on the ‘drinks per week’ GWAS^51^: 1) we selected single nucleotide polymorphisms (SNPs) present in HapMap phase 3^53,54^ and 1000 genomes phase 3^55^; 2) SNP weights were calculated with MegaPRS^56^ with the BLD-LDAK heritability model^57^, selecting the optimal PGS tool and hyperparameters (‘mega’ argument, for AUD: LDAK-Ridge with heritability of 0.70); and 3) individual-level PGSs were computed with PLINK (v2.00a5LM)^58,59^ and standardized (mean = 0, SD = 1) within each study.

### Statistical analyses

#### Survival analysis models

We performed Cox proportional hazard (Cox-PH) regression with age as the time scale. Follow-up began at birth and ended with the earliest disease diagnosis (cases), death, last registry follow-up, or age 80. For UK Biobank and Generation Scotland, we excluded diseases for which those cohorts contributed to the discovery GWAS; in FinnGen, overlapping discovery samples were excluded from analyses (**Supplementary Table 3**).

All models were adjusted for sex (except for breast and prostate cancer, which were respectively restricted to only females or only males), birth decade, and genetic principal components (PCs, 5 PCs in models without PGSs, 10 PCs in models with PGSs). Proportional hazards assumptions were tested in FinnGen with scaled Schoenfeld residuals (**Supplementary Fig.s 1-2**).

We evaluated four models for each disease in each study: 1) association of the disease-specific PGS or education alone with disease risk; 2) association of the disease-specific PGS and education together; 3) association of the disease-specific PGS stratified by education; and 4) association of the disease-specific PGS, education, and their interaction. By comparing models 1 and 2, we assessed the potential overlapping effects between genetic and educational risk factors. Because differences in the strength or direction of associations can occur even if the main effects are (largely) independent, models 3 and 4 evaluated whether interaction effects were present.

#### Meta-analyses

We conducted fixed-effects meta-analyses of log hazard ratios (i.e., beta coefficients) across cohorts with the ‘metafor’ (v. 4.6-0) R package (v4.4.1)^60,61^. Two exceptions were atrial fibrillation and colorectal cancer, where discovery GWAS overlap or small case counts precluded analysis in some cohorts, so results are based on FinnGen alone. To assess heterogeneity across studies we estimated Cochran’s *Q*-statistic^62^. To test for attenuation of PGS or education effects after mutual adjustment, we compared effect sizes using two-sided Wald tests. We applied a Bonferroni-adjusted significance threshold of *p* < 2.63×10^-03^ to account for 19 outcomes.

#### Cumulative incidence estimation

Cumulative incidence up to age 80 was estimated using age-, sex-, and country-specific incidence, prevalence, and mortality data from the GBD^18^, accounting for competing risk of death. Following previous approaches^19,63,64^, we applied a piecewise constant hazard model across 5-year age bins to convert incidence rates into probabilities of disease. These were combined with survival probabilities to estimate cumulative incidence.

For PGS-stratified incidence estimates, we used Cox-PH models in FinnGen to estimate HRs across five PGS groups (<20%, 20–40%, 40–60% [reference], 60–95%, >95%), stratified by education. These HRs were then applied to GBD-derived baseline rates. Confidence intervals were derived using bootstrapping, and age-specific HRs were interpolated across age quartiles as needed.

#### Evaluating predictive performance

In a random 80% training set of FinnGen participants, we trained five logistic regression models: 1) association of the basic covariates (2 models were run, both included sex [except for breast and prostate cancer], birth year and the first 5 genetic PCs, the second model also included genetic PCs 6-10) with disease risk; 2) association of the basic covariates and disease-specific PGS with disease risk 3) association of the basic covariates and educational attainment with disease risk; 4) association of the basic covariates, disease-specific PGS, and educational attainment; and 5) association of the basic covariates, disease-specific PGS, educational attainment, and their interaction. We evaluated performance of these five models in the remaining 20% of FinnGen and validate these results in the UK Biobank. Discrimination of the models was assessed with Area Under the Curve (AUC)^65^, continuous Net Reclassification Index (NRI), and continuous Integrated Discrimination Index (IDI)^66^. Here, we made three comparisons of the model performance: 1) compared models with basic covariates to models that also included the disease-specific PGS or educational attainment; 2) compared models with basic covariates, disease-specific PGS, and educational attainment to model that only included the basic covariates and the disease-specific PGS or educational attainment; and 3) compared model with basic covariates, disease-specific PGS, educational attainment, and their interaction with model that did not include the interaction between the disease-specific PGS and educational attainment.

The AUC quantifies a model’s ability to discriminate between cases and controls, where an AUC of 0.5 indicates no discriminative ability (i.e., equivalent to random chance), while an AUC of 1 indicates perfect discrimination. To compare the discriminative performance of the different models, we tested whether their AUCs were significantly different with DeLong’s test^67^. After assessing the differences in the AUC, calculated with the ‘pROC’ (v. 1.18.5) R package (v. 4.5.0)^68^, between the models in 20% of FinnGen, we validated these findings in the UK Biobank.

The NRI assesses whether a more complex model improves the risk classification compared to a less complex reference model. Thus, it measures how often individuals are correctly reclassified as cases or controls in both models. A positive NRI indicates the more complex model improves classification, while negative NRI estimates indicate the more complex model worsens classification as compared to the less complex reference model. In contrast, the IDI measures the overall improvement in risk prediction by comparing the difference in average predicted probabilities between cases and controls between two models. Here, a positive IDI indicates the more complex model has better discrimination ability than the less complex reference model, and a negative IDI indicates the more complex model is worse at distinguishing cases from controls than the less complex reference model. The NRI and IDI were calculated with the ‘nricens’ (v. 1.6) R package (v. 4.5.0)^69^.

Sensitivity analyses

### Fine-Gray competing risk models

For outcomes where a significant interaction between the disease-specific PGS and educational attainment was observed in the meta-analysis (any cancer, asthma, atrial fibrillation, coronary heart disease, hip osteoarthritis, knee osteoarthritis, and type 2 diabetes), we performed additional sensitivity analyses using Fine-Gray competing risk models in the FinnGen study^70^. In survival analysis, the presence of competing risks, events such as death from other causes that preclude the event of interest, can bias standard Cox-PH models by overestimating the cumulative incidences. The Fine-Gray model addresses this by directly modeling the subdistribution hazard, allowing for estimation of the cumulative incidence function of the disease outcome while accounting for the competing risk of death from other causes.

### Analyses in non-European ancestries

In UK Biobank, we explored whether the associations observed in individuals of European ancestry for the disease-specific PGS and educational attainment with type 1 diabetes, prostate cancer, gout, rheumatoid arthritis, breast cancer, epilepsy, and alcohol use disorder replicated in individuals of non-European ancestry (see **Supplementary Methods**). Given the relatively low sample sizes for non-European ancestries, we only had sufficient cases and controls by education level available for breast and prostate cancer in South Asian (SAS), East Asian, and African individuals, and for gout, rheumatoid arthritis, and epilepsy only in SAS individuals. All other trait-ancestry combinations could not be assessed.

### Analyses with occupation as socioeconomic index

To assess whether our results are robust across SES indices, we reran all Cox-PH models in FinnGen with occupation as alternative SES measure. We evaluated five models for each disease: 1) association of the disease-specific PGS or occupation alone with disease risk; 2) association of the disease-specific PGS and occupation together; 3) association of the disease-specific PGS stratified by level of occupation; 4) association of the disease-specific PGS, occupation, and their interaction; and 5) association of disease-specific PGS strata (<20%, 20–40%, 40–60% [reference], 60–95%, >95%), stratified by occupation. Model five was used to calculate cumulative incidences across PGS strata and occupation levels.

Information about occupation in FinnGen was retrieved from Statistics Finland using the classification of socioeconomic groups which in turn is based on the classification of occupations (for details see: https://stat.fi/en/luokitukset/sosioekon_asema/sosioekon_asema_1_19890101). Occupation was available for a maximum of 25 different years (1970-2020): 1970, 1975, 1980, 1990, 1993, 1995, 2000, and annually from 2004 onwards. Occupation was captured into 9 broad categories: 1) self-employed; 2) upper-level employees with administrative, managerial, professional and related occupations; 3) lower-level employees with administrative and clerical occupations; 4) manual workers; 5) students; 6) pensioners; 7) others (including unemployed); 8) unknown occupation; and 9) missing occupation. For each individual, we selected the occupation closest to the year of event for cases or end of follow-up for controls. In case of ties, i.e., when the event/end-of-follow-up fell in between measurements, we selected the earlier occupation information.

Whenever the occupation class closest to event/end-of-follow-up was ‘students’, ‘pensioners’, ‘other’, ‘unknown’, or ‘missing’, we iteratively extracted the occupation class at a previous census until an individual could be assigned to ‘self-employed’, ‘upper-level employees with administrative, managerial, professional and related occupations’, ‘lower-level employees with administrative and clerical occupations’, or ‘manuals workers’. Whenever this reassignment to an earlier census date was not possible, the individual was excluded from the analyses. To ensure easier comparison between the educational attainment and occupation analyses, we combined the ‘manual workers’ and ‘lower-level employees with administrative and clerical occupations’ into a single ‘lower-level’ occupation class and compared this group to individuals with ‘upper-level employees with administrative, managerial, professional and related occupations’, which we refer to as ‘upper-level’ occupation in the manuscript.

## Acknowledgements

We want to acknowledge the participants and investigators of the FinnGen study. Following biobanks are acknowledged for delivering biobank samples to FinnGen: Auria Biobank (www.auria.fi/biopankki), THL Biobank (www.thl.fi/biobank), Helsinki Biobank (www.helsinginbiopankki.fi), Biobank Borealis of Northern Finland (https://www.ppshp.fi/Tutkimus-ja-opetus/Biopankki/Pages/Biobank-Borealis-briefly-in-English.aspx), Finnish Clinical Biobank Tampere (www.tays.fi/en-US/Research_and_development/Finnish_Clinical_Biobank_Tampere), Biobank of Eastern Finland (www.ita-suomenbiopankki.fi/en), Central Finland Biobank (www.ksshp.fi/fi-FI/Potilaalle/Biopankki), Finnish Red Cross Blood Service Biobank (www.veripalvelu.fi/verenluovutus/biopankkitoiminta), Terveystalo Biobank (www.terveystalo.com/fi/Yritystietoa/Terveystalo-Biopankki/Biopankki/) and Arctic Biobank (https://www.oulu.fi/en/university/faculties-and-units/faculty-medicine/northern-finland-birth-cohorts-and-arctic-biobank). All Finnish Biobanks are members of BBMRI.fi infrastructure (https://www.bbmri-eric.eu/national-nodes/finland/). Finnish Biobank Cooperative -FINBB (https://finbb.fi/) is the coordinator of BBMRI-ERIC operations in Finland. We thank participants and scientists involved in making the UK Biobank resource available (http://www.ukbiobank.ac.uk/).

## Declarations

### Ethical approval

Study subjects in FinnGen provided (written) informed consent for biobank research, based on the Finnish Biobank Act. Alternatively, separate research cohorts, collected prior the Finnish Biobank Act came into effect (in September 2013) and start of FinnGen (August 2017), were collected based on study-specific consents and later transferred to the Finnish biobanks after approval by Fimea (Finnish Medicines Agency), the National Supervisory Authority for Welfare and Health. Recruitment protocols followed the biobank protocols approved by Fimea. The Coordinating Ethics Committee of the Hospital District of Helsinki and Uusimaa (HUS) statement number for the FinnGen study is Nr HUS/990/2017.

The FinnGen study is approved by Finnish Institute for Health and Welfare (permit numbers: THL/2031/6.02.00/2017, THL/1101/5.05.00/2017, THL/341/6.02.00/2018, THL/2222/6.02.00/2018, THL/283/6.02.00/2019, THL/1721/5.05.00/2019 and THL/1524/5.05.00/2020), Digital and population data service agency (permit numbers: VRK43431/2017-3, VRK/6909/2018-3, VRK/4415/2019-3), the Social Insurance Institution (permit numbers: KELA 58/522/2017, KELA 131/522/2018, KELA 70/522/2019, KELA 98/522/2019, KELA 134/522/2019, KELA 138/522/2019, KELA 2/522/2020, KELA 16/522/2020), Findata permit numbers THL/2364/14.02/2020, THL/4055/14.06.00/2020, THL/3433/14.06.00/2020, THL/4432/14.06/2020, THL/5189/14.06/2020, THL/5894/14.06.00/2020, THL/6619/14.06.00/2020, THL/209/14.06.00/2021, THL/688/14.06.00/2021, THL/1284/14.06.00/2021, THL/1965/14.06.00/2021, THL/5546/14.02.00/2020, THL/2658/14.06.00/2021, THL/4235/14.06.00/2021, Statistics Finland (permit numbers: TK-53-1041-17 and TK/143/07.03.00/2020 (earlier TK-53-90-20) TK/1735/07.03.00/2021, TK/3112/07.03.00/2021) and Finnish Registry for Kidney Diseases permission/extract from the meeting minutes on 4th July 2019.

The Biobank Access Decisions for FinnGen samples and data utilized in FinnGen Data Freeze 11 include: THL Biobank BB2017_55, BB2017_111, BB2018_19, BB_2018_34, BB_2018_67, BB2018_71, BB2019_7, BB2019_8, BB2019_26, BB2020_1, BB2021_65, Finnish Red Cross Blood Service Biobank 7.12.2017, Helsinki Biobank HUS/359/2017, HUS/248/2020, HUS/430/2021 §28, §29, HUS/150/2022 §12, §13, §14, §15, §16, §17, §18, §23, §58, §59, HUS/128/2023 §18, Auria Biobank AB17-5154 and amendment #1 (August 17 2020) and amendments BB_2021-0140, BB_2021-0156 (August 26 2021, Feb 2 2022), BB_2021-0169, BB_2021-0179, BB_2021-0161, AB20-5926 and amendment #1 (April 23 2020) and it’s modifications (Sep 22 2021), BB_2022-0262, BB_2022-0256, Biobank Borealis of Northern Finland_2017_1013, 2021_5010, 2021_5010 Amendment, 2021_5018, 2021_5018 Amendment, 2021_5015, 2021_5015 Amendment, 2021_5015 Amendment_2, 2021_5023, 2021_5023 Amendment, 2021_5023 Amendment_2, 2021_5017, 2021_5017 Amendment, 2022_6001, 2022_6001 Amendment, 2022_6006 Amendment, 2022_6006 Amendment, 2022_6006 Amendment_2, BB22-0067, 2022_0262, 2022_0262 Amendment, Biobank of Eastern Finland 1186/2018 and amendment 22§/2020, 53§/2021, 13§/2022, 14§/2022, 15§/2022, 27§/2022, 28§/2022, 29§/2022, 33§/2022, 35§/2022, 36§/2022, 37§/2022, 39§/2022, 7§/2023, 32§/2023, 33§/2023, 34§/2023, 35§/2023, 36§/2023, 37§/2023, 38§/2023, 39§/2023, 40§/2023, 41§/2023, Finnish Clinical Biobank Tampere MH0004 and amendments (21.02.2020 & 06.10.2020), BB2021-0140 8§/2021, 9§/2021, §9/2022, §10/2022, §12/2022, 13§/2022, §20/2022, §21/2022, §22/2022, §23/2022, 28§/2022, 29§/2022, 30§/2022, 31§/2022, 32§/2022, 38§/2022, 40§/2022, 42§/2022, 1§/2023, Central Finland Biobank 1-2017, BB_2021-0161, BB_2021-0169, BB_2021-0179, BB_2021-0170, BB_2022-0256, BB_2022-0262, BB22-0067, Decision allowing to continue data processing until 31st Aug 2024 for projects: BB_2021-0179, BB22-0067,BB_2022-0262, BB_2021-0170, BB_2021-0164, BB_2021-0161, and BB_2021-0169, and Terveystalo Biobank STB 2018001 and amendment 25th Aug 2020, Finnish Hematological Registry and Clinical Biobank decision 18th June 2021, Arctic biobank P0844: ARC_2021_1001.

Ethics approval for the UK Biobank study was obtained from the North West Centre for Research Ethics Committee (11/NW/0382). UK Biobank data used in this study were obtained under approved application 78537. Ethical approval for the GS:SFHS study was obtained from the Tayside Committee on Medical Research Ethics (on behalf of the National Health Service; REC Reference Number: 05/S1401/89).

### Data availability

The raw individual-level data are protected and are not publicly available due to data privacy laws, but they can be accessed through individual participating biobanks. The Finnish biobank data can be accessed through the Fingenious® services (https://site.fingenious.fi/en/) managed by FINBB. UK Biobank data are available through a procedure described at http://www.ukbiobank.ac. GS data may be accessed through an application process described here: https://www.ed.ac.uk/generationscotland/for-researchers/access. The GWAS data used in this study are available in the GWAS catalog database under accession codes listed in **Supplementary Table 2**. The PGS scores used in this study are available in the PGS Catalog under publication ID: PGP000618 and score IDs: PGS004869-PGS004886. All other data generated during this study are included in this published article and its supplementary information files.

### Code availability

The code used for these analyses is available at https://github.com/intervene-EU-H2020/GxE_SESDisease

### Author contributions

Conceptualization: P.M., N.M. and S.R., Data curation: M.T., K.D., Z.Y. and T.H., Formal Analysis: F.A.H. and A.R., Funding acquisition: A.G. and S.R., Project administration: F.A.H., Supervision: S.R., Validation: F.A.H., A.R., D.L.M. and R.E.M., Visualization: F.A.H., M.T. and K.D. Writing – original draft: F.A.H. and S.R. Writing – review & editing: A.R., M.T., K.D., T.H., D.L.M., R.E.M., P.M. and N.M.

## References

1. Dalstra, J. et al. Socioeconomic differences in the prevalence of common chronic diseases: an overview of eight European countries. Int. J. Epidemiol. 34, 316–326 (2005).

2. Kivimäki, M. et al. Association between socioeconomic status and the development of mental and physical health conditions in adulthood: a multi-cohort study. Lancet Public Health 5, e140–e149 (2020).

3. Hvidberg, M. F., Frølich, A. & Lundstrøm, S. L. Catalogue of socioeconomic disparities and characteristics of 199+ chronic conditions—A nationwide register-based population study. PLOS ONE 17, e0278380 (2022).

4. Lundqvist, A., Andersson, E., Ahlberg, I., Nilbert, M. & Gerdtham, U. Socioeconomic inequalities in breast cancer incidence and mortality in Europe—a systematic review and meta-analysis. Eur. J. Public Health 26, 804–813 (2016).

5. Alfonso, J. H. et al. Occupation and cutaneous melanoma: a 45-year historical cohort study of 14·9 million people in five Nordic countries*. Br. J. Dermatol. 184, 672–680 (2021).

6. Abdellaoui, A., Yengo, L., Verweij, K. J. H. & Visscher, P. M. 15 years of GWAS discovery: Realizing the promise. Am. J. Hum. Genet. 110, 179–194 (2023).

7. Wray, N. R., Goddard, M. E. & Visscher, P. M. Prediction of individual genetic risk to disease from genome-wide association studies. Genome Res. 17, 1520–1528 (2007).

8. Kullo, I. J. et al. Polygenic scores in biomedical research. Nat. Rev. Genet. 23, 524–532 (2022).

9. Mars, N. et al. Genome-wide risk prediction of common diseases across ancestries in one million people. Cell Genomics 2, (2022).

10. Sun, T.-H. et al. Utility of polygenic scores across diverse diseases in a hospital cohort for predictive modeling. Nat. Commun. 15, 3168 (2024).

11. Elliott, J. et al. Predictive Accuracy of a Polygenic Risk Score–Enhanced Prediction Model vs a Clinical Risk Score for Coronary Artery Disease. JAMA 323, 636–645 (2020).

12. Mars, N. et al. Polygenic and clinical risk scores and their impact on age at onset and prediction of cardiometabolic diseases and common cancers. Nat. Med. 26, 549–557 (2020).

13. Mars, N. et al. Systematic comparison of family history and polygenic risk across 24 common diseases. Am. J. Hum. Genet. 109, 2152–2162 (2022).

14. Okbay, A. et al. Polygenic prediction of educational attainment within and between families from genome-wide association analyses in 3 million individuals. Nat. Genet. 54, 437–449 (2022).

15. Dar-Nimrod, I. & Heine, S. J. Genetic Essentialism: On the Deceptive Determinism of DNA. Psychol. Bull. 137, 800–818 (2011).

16. Liu, J. et al. Polygenic Risk Score, Lifestyles, and Type 2 Diabetes Risk: A Prospective Chinese Cohort Study. Nutrients 15, 2144 (2023).

17. Khera, A. V. et al. Genetic Risk, Adherence to a Healthy Lifestyle, and Coronary Disease. N. Engl. J. Med. 375, 2349–2358 (2016).

18. Vos, T. et al. Global burden of 369 diseases and injuries in 204 countries and territories, 1990–2019: a systematic analysis for the Global Burden of Disease Study 2019. The Lancet 396, 1204–1222 (2020).

19. Jermy, B. et al. A unified framework for estimating country-specific cumulative incidence for 18 diseases stratified by polygenic risk. Nat. Commun. 15, 5007 (2024).

20. Stringhini, S. et al. Socioeconomic status and the 25 × 25 risk factors as determinants of premature mortality: a multicohort study and meta-analysis of 1·7 million men and women. The Lancet 389, 1229–1237 (2017).

21. Clark, A. M., DesMeules, M., Luo, W., Duncan, A. S. & Wielgosz, A. Socioeconomic status and cardiovascular disease: risks and implications for care. Nat. Rev. Cardiol. 6, 712–722 (2009).

22. Zhou, L. et al. Exploring socioeconomic status, lifestyle factors, and cardiometabolic disease outcomes in the United States: insights from a population-based cross-sectional study. BMC Public Health 24, 2174 (2024).

23. Judd, J., Spence, J. P., Pritchard, J. K., Kachuri, L. & Witte, J. S. Investigating the role of neighborhood socioeconomic status and germline genetics on prostate cancer risk. Hum. Genet. Genomics Adv. 6, (2025).

24. Chetty, R. et al. The Association Between Income and Life Expectancy in the United States, 2001-2014. JAMA 315, 1750–1766 (2016).

25. White, M. C. et al. Age and Cancer Risk: A Potentially Modifiable Relationship. Am. J. Prev. Med. 46, S7 (2014).

26. Bozhar, H. et al. Socio-economic inequality of utilization of cancer testing in Europe: A cross-sectional study. Prev. Med. Rep. 26, 101733 (2022).

27. Martikainen, P. et al. Joint association between education and polygenic risk score for incident coronary heart disease events: a longitudinal population-based study of 26 203 men and women. J Epidemiol Community Health 75, 651–657 (2021).

28. Carter, A. R. et al. Educational attainment as a modifier for the effect of polygenic scores for cardiovascular risk factors: cross-sectional and prospective analysis of UK Biobank. Int. J. Epidemiol. 51, 885–897 (2022).

29. Abdellaoui, A. et al. Socio-economic status is a social construct with heritable components and genetic consequences. *Nat*. Hum. Behav. 1–13 (2025) doi:10.1038/s41562-025-02150-4.

30. Evans, G. W. & English, K. The Environment of Poverty: Multiple Stressor Exposure, Psychophysiological Stress, and Socioemotional Adjustment. Child Dev. 73, 1238–1248 (2002).

31. Kraft, P. & Kraft, B. Explaining socioeconomic disparities in health behaviours: A review of biopsychological pathways involving stress and inflammation. Neurosci. Biobehav. Rev. 127, 689– 708 (2021).

32. Cutler, D. M. & Lleras-Muney, A. Understanding Differences in Health Behaviors by Education. J. Health Econ. 29, 1 (2009).

33. Chen, E. & Miller, G. E. Socioeconomic Status and Health: Mediating and Moderating Factors. Annu. Rev. Clin. Psychol. 9, 723–749 (2013).

34. Wood, A. M. et al. Risk thresholds for alcohol consumption: combined analysis of individual-participant data for 599 912 current drinkers in 83 prospective studies. The Lancet 391, 1513–1523 (2018).

35. Khan, T. A. et al. Combination of Multiple Low-Risk Lifestyle Behaviors and Incident Type 2 Diabetes: A Systematic Review and Dose-Response Meta-analysis of Prospective Cohort Studies. Diabetes Care 46, 643–656 (2023).

36. Svendsen, M. T. et al. Associations of health literacy with socioeconomic position, health risk behavior, and health status: a large national population-based survey among Danish adults. BMC Public Health 20, 565 (2020).

37. Berete, F. et al. Does health literacy mediate the relationship between socioeconomic status and health related outcomes in the Belgian adult population? BMC Public Health 24, 1182 (2024).

38. Martin, A. R. et al. Clinical use of current polygenic risk scores may exacerbate health disparities. Nat. Genet. 51, 584–591 (2019).

39. Siddiq, H. & Najand, B. Immigration Status, Socioeconomic Status, and Self-Rated Health in Europe. Int. J. Environ. Res. Public. Health 19, 15657 (2022).

40. OECD. Finland: Country Health Profile 2023. (Organisation for Economic Co-operation and Development, Paris, 2023).

41. OECD. United Kingdom: Country Health Profile 2019. (Organisation for Economic Co-operation and Development, Paris, 2019).

42. OECD. Education at a Glance 2021: OECD Indicators. (Organisation for Economic Co-operation and Development, Paris, 2021).

43. Dow, W. H., Schoeni, R. F., Adler, N. E. & Stewart, J. Evaluating the evidence base: Policies and interventions to address socioeconomic status gradients in health. Ann. N. Y. Acad. Sci. 1186, 240–251 (2010).

44. Moore, G. F., Littlecott, H. J., Turley, R., Waters, E. & Murphy, S. Socioeconomic gradients in the effects of universal school-based health behaviour interventions: a systematic review of intervention studies. BMC Public Health 15, 907 (2015).

45. International Standard Classification of Education, ISCED 1997. in Advances in Cross-National Comparison: A European Working Book for Demographic and Socio-Economic Variables (eds. Hoffmeyer-Zlotnik, J. H. P. & Wolf, C.) 195–220 (Springer US, Boston, MA, 2003). doi:10.1007/978-1-4419-9186-7_10.

46. Galobardes, B., Shaw, M., Lawlor, D. A., Lynch, J. W. & Smith, G. D. Indicators of socioeconomic position (part 1). J. Epidemiol. Community Health 60, 7–12 (2006).

47. Kurki, M. I. et al. FinnGen provides genetic insights from a well-phenotyped isolated population. Nature 613, 508–518 (2023).

48. Bycroft, C. et al. The UK Biobank resource with deep phenotyping and genomic data. Nature 562, 203–209 (2018).

49. Smith, B. H., et al. Cohort Profile: Generation Scotland: Scottish Family Health Study (GS:SFHS). The study, its participants and their potential for genetic research on health and illness. Int. J. Epidemiol. 42, 689–700 (2013).

50. Collins, S. E. Associations Between Socioeconomic Factors and Alcohol Outcomes. Alcohol Res. Curr. Rev. 38, 83–94 (2016).

51. Saunders, G. R. B. et al. Genetic diversity fuels gene discovery for tobacco and alcohol use. Nature 612, 720–724 (2022).

52. Lambert, S. A. et al. The Polygenic Score Catalog: new functionality and tools to enable FAIR research. 2024.05.29.24307783 Preprint at 10.1101/2024.05.29.24307783 (2024).

53. Gibbs, R. A. et al. The International HapMap Project. Nature 426, 789–796 (2003).

54. Altshuler, D., Donnelly, P., & The International HapMap Consortium. A haplotype map of the human genome. Nature 437, 1299–1320 (2005).

55. Auton, A. et al. A global reference for human genetic variation. Nature 526, 68–74 (2015).

56. Zhang, Q., Privé, F., Vilhjálmsson, B. & Speed, D. Improved genetic prediction of complex traits from individual-level data or summary statistics. Nat. Commun. 12, 4192 (2021).

57. Speed, D., Holmes, J. & Balding, D. J. Evaluating and improving heritability models using summary statistics. Nat. Genet. 52, 458–462 (2020).

58. Purcell, S. et al. PLINK: A Tool Set for Whole-Genome Association and Population-Based Linkage Analyses. Am. J. Hum. Genet. 81, 559–575 (2007).

59. Chang, C. C. et al. Second-generation PLINK: rising to the challenge of larger and richer datasets. GigaScience 4, s13742-015-0047–8 (2015).

60. Viechtbauer, W. Conducting Meta-Analyses in R with the metafor Package. J. Stat. Softw. 36, 1–48 (2010).

61. R Core Team. R: A Language and Environment for Statistical Computing. R Foundation for Statistical Computing (2024).

62. Cochran, W. G. The Combination of Estimates from Different Experiments. Bioethics (1954) doi:10.2307/3001666.

63. The GBD 2016 Lifetime Risk of Stroke Collaborators. Global, Regional, and Country-Specific Lifetime Risks of Stroke, 1990 and 2016. N. Engl. J. Med. 379, 2429–2437 (2018).

64. Jukarainen, S. et al. Genetic risk factors have a substantial impact on healthy life years. Nat. Med. 28, 1893–1901 (2022).

65. Hanley, J. A. & McNeil, B. J. The meaning and use of the area under a receiver operating characteristic (ROC) curve. Radiology 143, 29–36 (1982).

66. Pencina, M. J., D’ Agostino Sr, R. B., D’ Agostino Jr, R. B. & Vasan, R. S. Evaluating the added predictive ability of a new marker: From area under the ROC curve to reclassification and beyond. Stat. Med. 27, 157–172 (2008).

67. DeLong, E. R., DeLong, D. M. & Clarke-Pearson, D. L. Comparing the Areas under Two or More Correlated Receiver Operating Characteristic Curves: A Nonparametric Approach. Biometrics 44, 837–845 (1988).

68. Robin, X. et al. pROC: an open-source package for R and S+ to analyze and compare ROC curves. BMC Bioinformatics 12, 77 (2011).

69. Inoue, E. nricens: NRI for Risk Prediction Models with Time to Event and Binary Response Data. (2018).

70. Fine, J. P. & and Gray, R. J. A Proportional Hazards Model for the Subdistribution of a Competing Risk. J. Am. Stat. Assoc. 94, 496–509 (1999).

