## Supplementary Methods and Figures for "The impact of polygenic score and socioeconomic status in predicting risk for 19 complex diseases"

#### **Study specific quality control**

##### **FinnGen (Data Freeze 11)**

###### **Registry data**

The construction of phenotype data in FinnGen involves the aggregation of information from nationwide electronic health registers. This encompasses nearly all aspects of a patient's interactions with the health service, including hospitalizations, medications, procedures, and deaths. The project's 18 registers are presented below, organized by their follow-up times.

- [Finnish Cancer Registry](https://cancerregistry.fi/) - From 1953
- [Register of Congenital Malformations](https://thl.fi/en/web/thlfi-en/statistics-and-data/data-and-services/register-descriptions/register-of-congenital-malformations) - From 1963
- [Reimbursement](https://raportit.kela.fi/ibi_apps/WFServlet?IBIF_ex=NIT137AL&YKIELI=E) - From 1964
- [Population Register](https://dvv.fi/en/population-information-system) - From 1964
- [Finnish Registry for Kidney Diseases](https://www.muma.fi/liitto/suomen_munuaistautirekisteri/finnish_registry_for_kidney_diseases) - From 1964
- [Causes of Death](https://www.stat.fi/til/ksyyt/index_en.html) - From 1969
- [Care Register for Health Care Inpatient Visits, HILMO](https://thl.fi/en/web/thlfi-en/statistics-and-data/data-and-services/register-descriptions/care-register-for-health-care) - From 1969
- [Socio-economic data](https://taika.stat.fi/en/) - From 1970
- [The Finnish Registry of Visual Impairment](https://www.nkl.fi/en) - From 1983
- [Medical Birth Register](https://thl.fi/en/web/thlfi-en/statistics-and-data/data-and-services/register-descriptions/newborns) - From 1987
- [Finnish National Infectious Disease Register](https://thl.fi/en/web/infectious-diseases-and-vaccinations/surveillance-and-registers/finnish-national-infectious-diseases-register) - From 1989
- [Cervical Cancer Screening](https://cancerregistry.fi/screening/cervical-cancer-screening/) - From 1991
- [Breast Cancer Screening](https://cancerregistry.fi/screening/breast-cancer-screening/) - From 1992
- [Drug Purchases](https://www.kela.fi/kelas-research-and-statistics) - From 1995
- [The Care Register for Social Welfare](https://www.julkari.fi/bitstream/handle/10024/127104/Tr21_15.pdf?sequence=4&isAllowed=y) - From 1995
- [Care Register for Health Care, specialist outpatient visits, HILMO](https://thl.fi/en/web/thlfi-en/statistics-and-data/data-and-services/register-descriptions/care-register-for-health-care) - From 1998
- [Register of Primary Health Care Visits, Avohilmo](https://thl.fi/fi/tilastot-ja-data/ohjeet-tietojen-toimittamiseen/perusterveydenhuollon-avohoidon-hoitoilmoitus-avohilmo) - From 2011
- [The Finnish Vaccination Register](https://thl.fi/en/web/infectious-diseases-and-vaccinations/surveillance-and-registers/finnish-national-vaccination-register-and-monitoring-of-the-vaccination-programme) - From 2011

Note: Although primary health care visits are covered in FinnGen, they are typically not included in the endpoints. Therefore, we only analyze secondary care data for our disease outcomes.

###### **Genotyping and quality control**

In FinnGen, there are prospectively recruited samples as well as legacy cohorts with pre-existing genotype data^1^. Genotyping of prospective samples was done with the ThermoFisher Axiom custom array (*N* = 371,914), which covers 655,973 variants. Genotype calling was conducted with the Array Power Tools software (<https://www.thermofisher.com/us/en/home/life-science/microarray-analysis/microarray-analysis-partners-programs/affymetrix-developers-network/affymetrix-power-tools.html>). Various Illumina arrays were used to genotype legacy cohorts (*N* = 81,819, for details see Kurki et al. (2023)^1^), and either GenCall or zCall algorithms were employed for genotype calling^2^.

The same quality control (QC) metrics were applied to both prospective and legacy cohorts. QC was performed per genotyping batch, information on the number of samples and variants removed per batch is provided in **Supplementary Table 35**.

Samples were removed if:

- The proportion of identity-by-decent (PLINK’s v2^3^ Pi hat) was > 0.9 and the samples were not monozygotic or replicates
- The reported sex did not match the genetically determined sex (F value ≤ 0.3 for females and ≥ 0.8 for males).
- Missingness was ≥ 5%
- Heterozygosity was ±4 standard deviations from the population average
- Pi hat was > 0.1 with 14 or more samples
- The samples deviated by ±4 standard deviations from the population average based on the first two genetic principal components.

Samples were tagged should there be evidence of a mendelian error or contain replicate samples with over 50,000 discrepancies.

Variants were removed if:

- The variant did not pass the Hardy-Weinberg Equilibrium test (p-value < 10^-6^).
- The variant had a call rate < 98%

###### **Imputation**

Pre-phasing was done with Eagle 2.4.1^4^, and imputation of samples was carried out using the SiSu v4.2 imputation reference panel^5^. The reference panel is designed specifically for the Finnish population, consisting of high-coverage (25-30x) whole-genome sequencing data from 8,554 Finns and 20,175,454 variants with a minor allele count of at least 3. After imputation, 9,641,808 variants were imputed with good quality (INFO > 0.6).

###### **Ancestry assignment**

First, the FinnGen samples were integrated with the 1000 genomes phase 3 dataset^6^. Genetic principal components were calculated using a subset of 180,042 pruned SNPs. Aberrant was used to identify and remove samples that deviated from the main cluster (*N* = 17,133)^7^. A probability of belonging to either a North-Western European or Finnish population was calculated by firstly performing Principal Component Analysis (PCA) in PLINK v2^3^ with individuals belonging to these ancestries from 1000 genomes data. FinnGen samples were then projected onto this PCA space and Mahalanobis distances calculated for each sample against each of the two ancestries. Samples were retained if there was ≥ 95% probability of belonging to the Finnish ancestry cluster (22 individuals removed).

##### **UK Biobank**

###### **Registry data**

The relevant columns used to define the phenotypes were:

- Cause of Death Primary (Column ID: 40001)
- Cause of Death Secondary (Column ID: 40002)
- Summary ICD10 Diagnoses (Column ID: 41270)
- Summary ICD10 Diagnoses Date (Column ID: 41280)
- Summary ICD9 Diagnoses (Column ID: 41271)
- Summary ICD9 Diagnoses Date (Column ID: 41281)

Such data are taken from the hospital episode statistics which relate to hospital inpatient data. For more information, please see <https://biobank.ndph.ox.ac.uk/showcase/showcase/docs/HospitalEpisodeStatistics.pdf>. Registry coverage depends on the country with follow-up beginning in 1997, 1998 and 1981 for England, Wales and Scotland respectively. End of follow-up was stated as 31st January 2021.

###### **Genotyping and quality control**

Two arrays were used to genotype UK Biobank participants^8^. The UK Biobank Lung Exome Variant Evaluation (UKBiLEVE) Axiom array was used to genotype 49,950 participants. The remaining 438,427 participants were genotypes using the Applied Biosystems UK Biobank Axiom Array.

PCA (fastPCA^9^) was performed on the genetic data and centralized QC on variants was performed on individuals identified to belong to the largest cluster (N=463,844) according to Aberrant - an unsupervised clustering algorithm^7^. Variants were assessed for evidence of allele frequency variation across batch, plate, sex or array and that genotypes were largely consistent with Hardy-Weinberg Equilibrium expectations (all *p*-value thresholds < 10^-12^). If a variant failed one or more tests within a given batch, it was set to missing. Previous research provides more detailed information on testing^8^.

###### **Imputation**

For 487,442 individuals, imputation was performed using the IMPUTE4 software^10^. Genetic variation from the Haplotype Reference Consortium (HRC)^11^ and merged UK10K+1000 Genomes were used as a reference panel^12^. Single Nucleotide Polymorphisms (SNPs) were only included in the final imputation if they were present in both reference panels, giving a total of 96,959,328 SNPs.

###### **Ancestry assignment**

Ancestry assignment uses methodology and scripts from GenoPred (https://opain.github.io/GenoPred/DiverseAncestry.html)^13^. Individuals were stratified into one of five super populations African (AFR), American (AMR), South Asian (SAS), East Asian (EAS) and European (EUR). The 1000 Genomes data acted as a reference given the individuals are known to belong to one of the 5 super populations^6^. Only unambiguous SNPs also present in both the HapMap3 consortium^14^ and the imputed UK Biobank data were retained for PCA. SNPs within both the reference (1000 Genomes) and target (UK Biobank) samples underwent quality control such that the minor allele frequency (MAF) > 5% (97,172 SNPs removed), variant missingness > 2% (57,916 SNPs removed), and Hardy-Weinberg Equilibrium p-value > 1e^-6^ (126,195 SNPs removed). 467,970 autosomal SNPs remained following QC and were in the intersection of the reference and target samples. Regions with long range linkage disequilibrium (e.g., inversions, for details see^8,15^) were excluded and independent SNPs (SNPs greater than 1000kb apart and r^2^ < 0.2) retained. PCA was then performed in the reference sample using PLINK v2^3^ and a multinomial elastic-net regression was trained using 5-fold cross validation, super population as the outcome and the first 10 PCs as covariates. PCs from the target sample were then projected into the reference space and prediction on super population made. Classifications were made according to the super population with the greatest probability. To be classified the max probability must be over 0.5, otherwise it was set to missing (*N* = 4,899).

PCA was performed using a random subset of 1000 individuals per super population and PC’s from the rest of the super population sample projected onto this space. Distances from the centroid were calculated and outliers removed. Outliers were classified as having a distance > 75 percentile + 30*Interquartile Range. Following within-ancestry QC, 8,414; 1,177; 2,951; 458,715 and 11,247 individuals were allocated to AFR, AMR, EAS, EUR and SAS super populations respectively.

##### **Generation Scotland**

###### **Registry data**

Disease outcomes were ascertained through linkage to primary (GP) and secondary (hospital) healthcare records. Individuals were subsetted to those registered at a GP that consented to sharing of primary records. GP records consisted of Read2 codes, which were mapped to ICD-10. Hospital data were obtained from Scottish Morbidity Records (SMR) where disease outcomes were coded using ICD-9 (pre March 1997) or ICD-10 (post March 1997). Start of follow-up was considered to be the latest date between Date of birth or March 1980, the date of GP linkage. End of follow up was considered to be October 2020 (the date to which GP data is available), date of death, or date of disease onset if after October 2020 (hospital data is available until March 2022).

###### **Genotyping and quality control**

Generation Scotland (GS) consists of ~24,000 individuals from across Scotland aged between 18-99 years. Phenotypic data were obtained at baseline along with whole blood samples for DNA quantification.

Genotype data was assayed for 20,195 participants in two batches with 9,863 participants in the first batch and the remainder in the second. The genotyping was performed using the Illumina HumanOmniExpressExome-8 v1.0 BeadChip and the Illumina HumanOmniExpressExome-8 v1.2 BeadChip, respectively, comprising 936,957 variants prior to QC. Individuals or SNPs with a low call rate (<98%) and SNPs with Hardy-Weinberg *p*-value<1x10^-6^ were removed. Mendelian errors were removed by setting the individual-level genotypes at erroneous SNPs to missing. In total, 163 samples and 332,099 variants were removed during QC, resulting in 604,858 variants prior to imputation.

###### **Imputation**

Genotyped data were imputed using the HRC panel v1^11^ with the positional Burrows-Wheeler transform software (v. 3)^16^ on the Sanger server. Autosomal haplotypes were checked to ensure consistency with the reference panel (strand orientation, reference allele, position). Pre-phasing was performed using Shapeit2 v2r837^17,18^ using the Shapeit2 duohmm option^19^ and cohort family structure in order to improve imputation quality^16^. Variants with low imputation quality (INFO<0.4) as well as monogenic variants were removed from the imputed set resulting in 24,111,857 variants for downstream analysis.

###### **Ancestry assignment**

Ancestry outliers were removed from the dataset. These were defined as individuals who were more than six standard deviations away from the mean in a principal component analysis, as performed with ACTA (v. 0.9)^20^, of GS merged with 1092 participants from the 1000 Genomes Project^6^.

### **Supplementary Figures**

**Supplementary Fig. 1.** Scaled Schoenfeld residuals of Cox proportional hazard models including the disease-specific polygenic score and educational attainment in the FinnGen study.

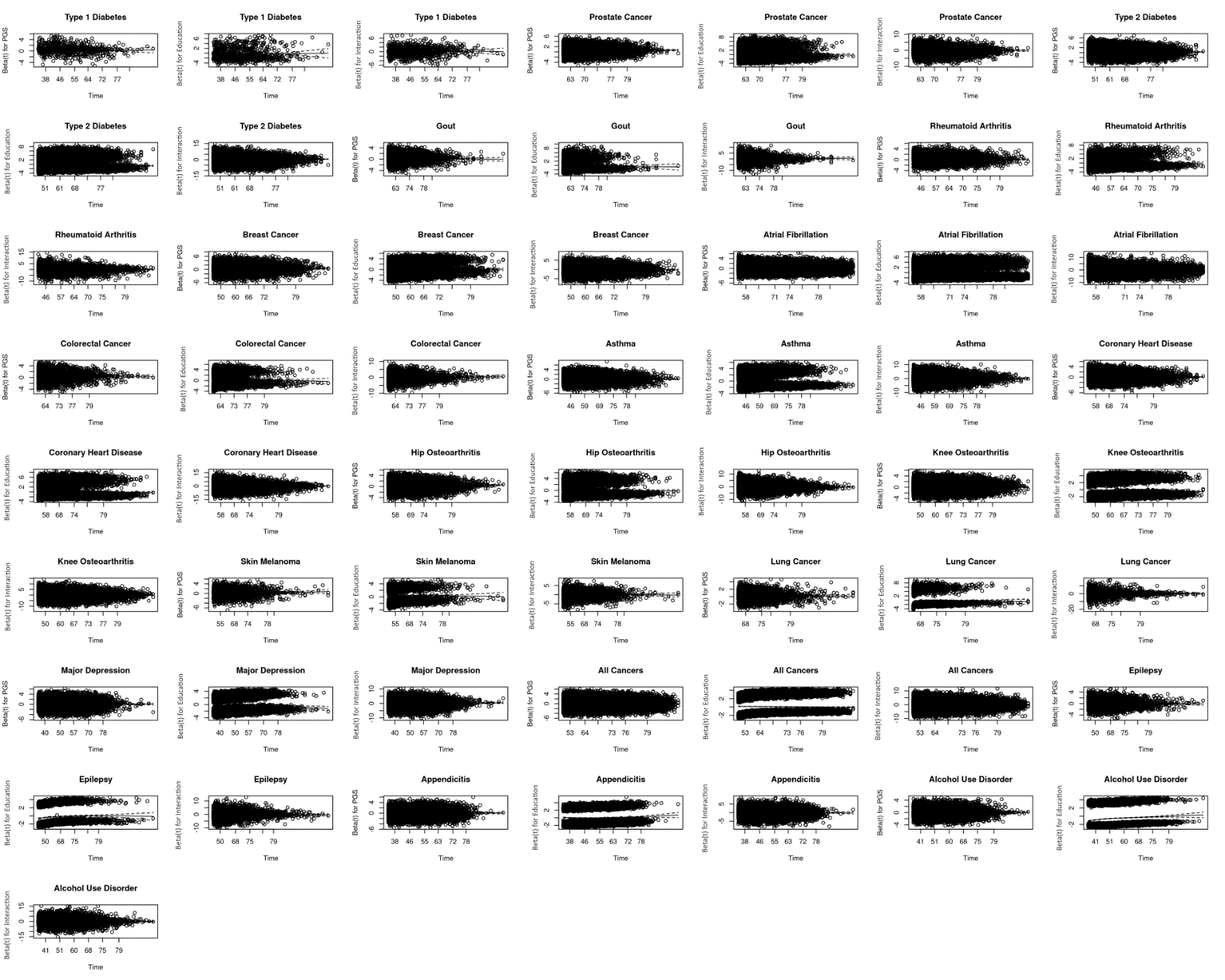

**Supplementary Fig. 2.** Scaled Schoenfeld residuals of Cox proportional hazard models including the disease-specific polygenic score, educational attainment, and their interaction in the FinnGen study.

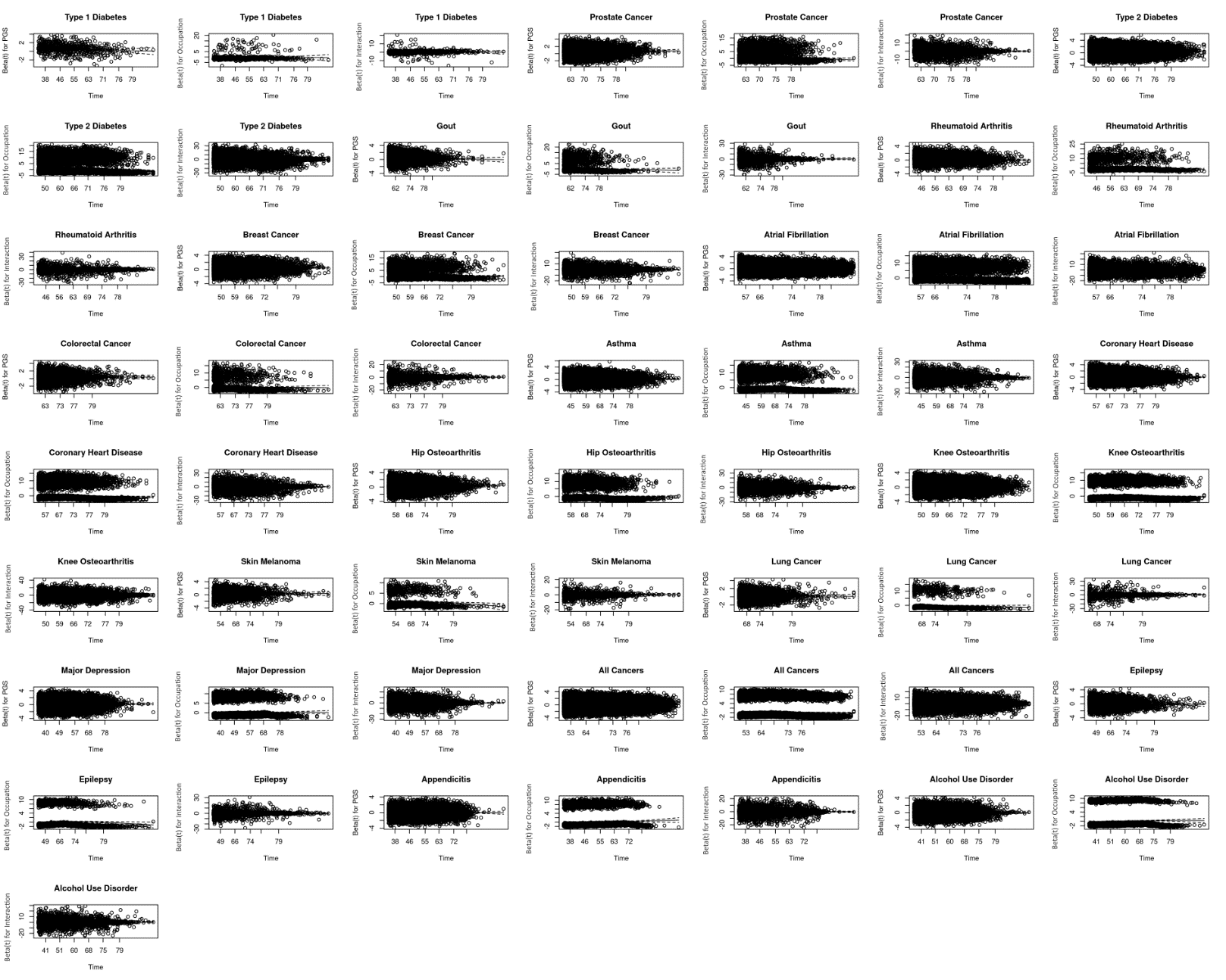

**Supplementary Fig. 3.** Meta-analyzed and biobank study-specific hazard ratios per standard deviation of the disease-specific polygenic scores (black) and for individuals with high education relative to individuals with low education (purple) on complex disease risk where the disease-specific polygenic scores and education were modelled separately. The asterisk indicates the complex disease was not meta-analyzed and assessed only in the FinnGen study (see **Methods** for details). The exact values are in **Supplementary Tables 5**. Sample sizes by case-control status and educational attainment for each complex disease in each biobank study are in **Supplementary Table 4**.

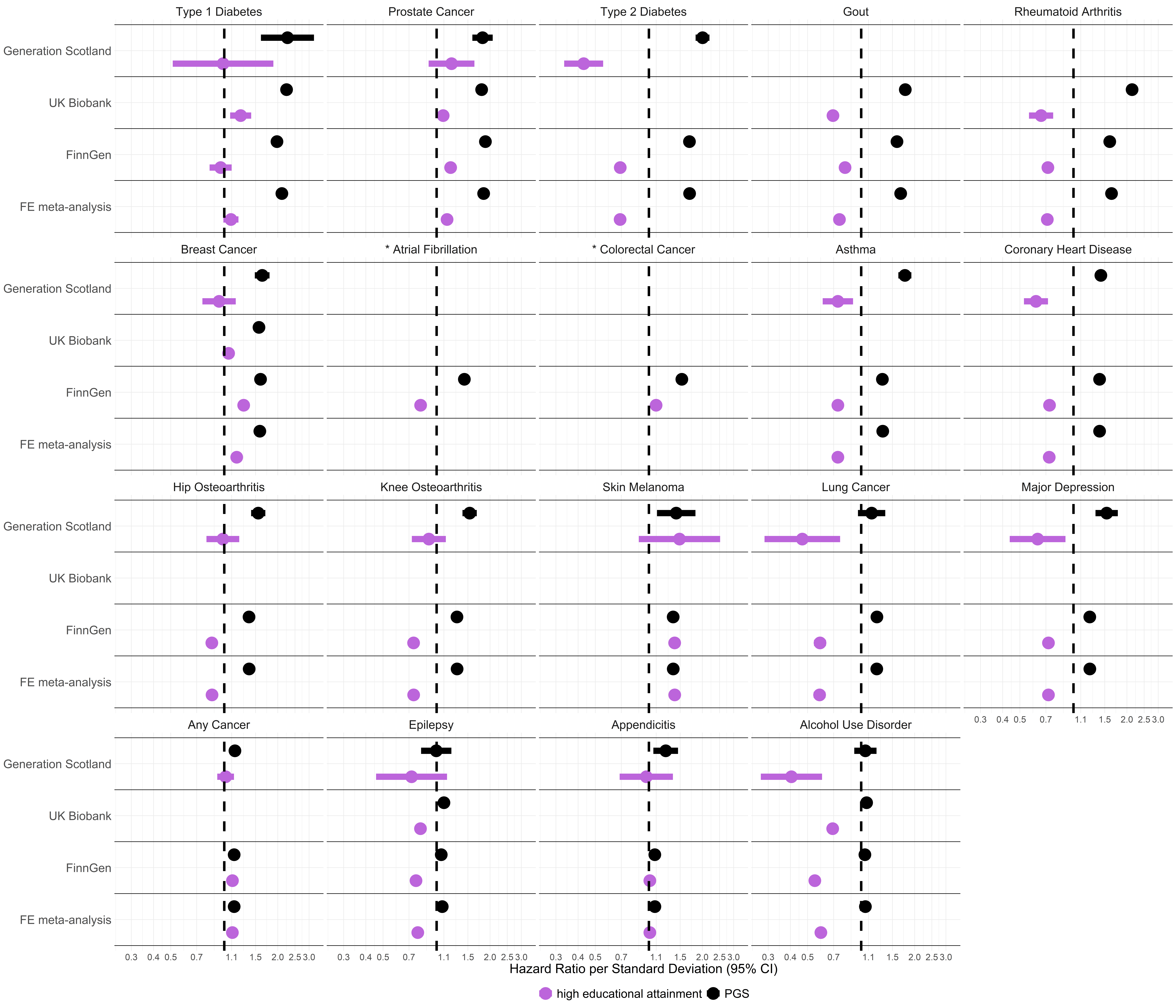

**Supplementary Fig. 4.** Comparison of the hazard ratio per standard deviation of the disease-specific polygenic scores on complex disease risk unadjusted (x-axis) and adjusted (y-axis) for the effect of high educational attainment (**A**) and comparison of the hazard ratio for individuals with high education relative to individuals with low education on complex disease risk unadjusted (x-axis) and adjusted (y-axis) for the disease-specific polygenic score (**B**)**.** The asterisk indicates the complex disease was not meta-analyzed and assessed only in the FinnGen study (see **Methods** for details). The exact values are in **Supplementary Tables 5-7**. Sample sizes by case-control status and educational attainment for each complex disease in each biobank study are in **Supplementary Table 4**.

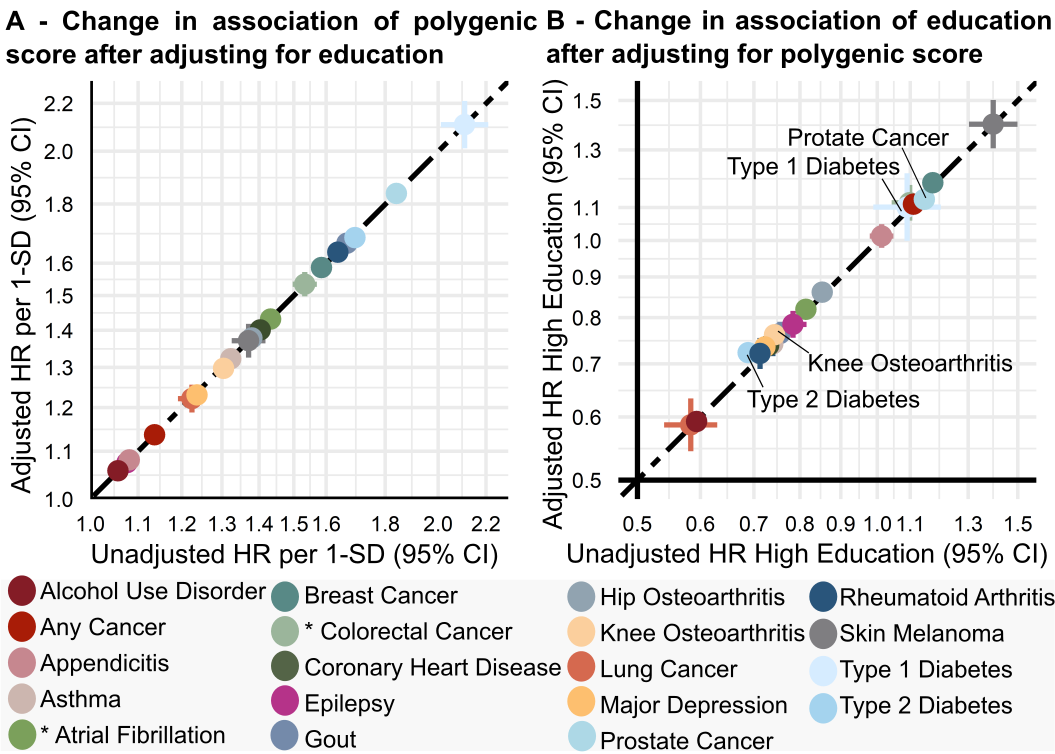

**Supplementary Fig. 5.** Meta-analyzed and biobank study-specific hazard ratios per standard deviation of the disease-specific polygenic scores (black) and for individuals with high education relative to individuals with low education (purple) on complex disease risk where the disease-specific polygenic scores and education were modelled jointly. The asterisk indicates the complex disease was not meta-analyzed and assessed only in the FinnGen study (see **Methods** for details). The exact values are in **Supplementary Table 6**. Sample sizes by case-control status and educational attainment for each complex disease in each biobank study are in **Supplementary Table 4**.

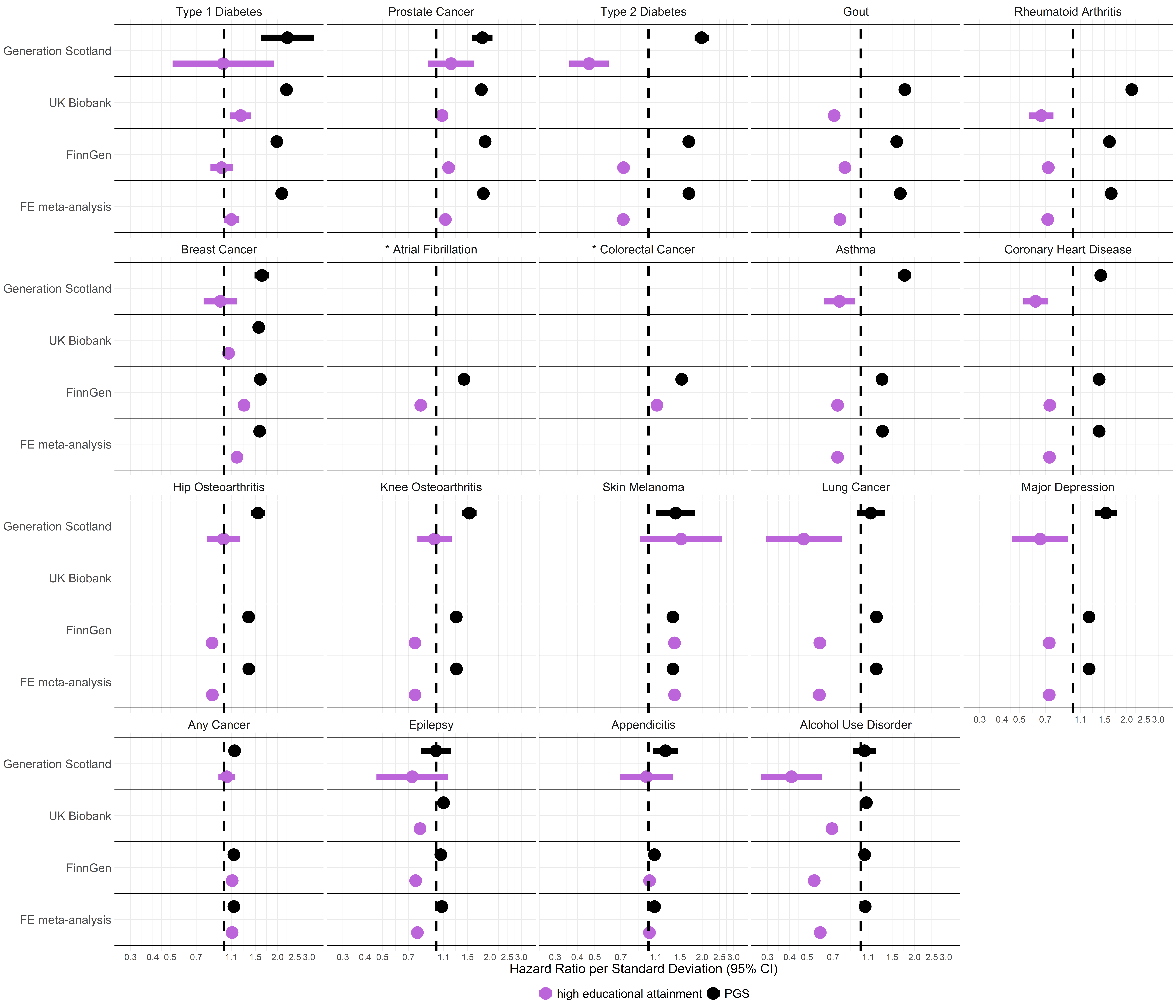

**Supplementary Fig. 6.** Meta-analyzed and biobank study-specific hazard ratios per standard deviation of the disease-specific polygenic scores by educational attainment on complex disease risk. The asterisk indicates the complex disease was not meta-analyzed and assessed only in the FinnGen study (see **Methods** for details). The exact values are in **Supplementary Table 8**. Sample sizes by case-control status and educational attainment for each complex disease in each biobank are in **Supplementary Table 4**.

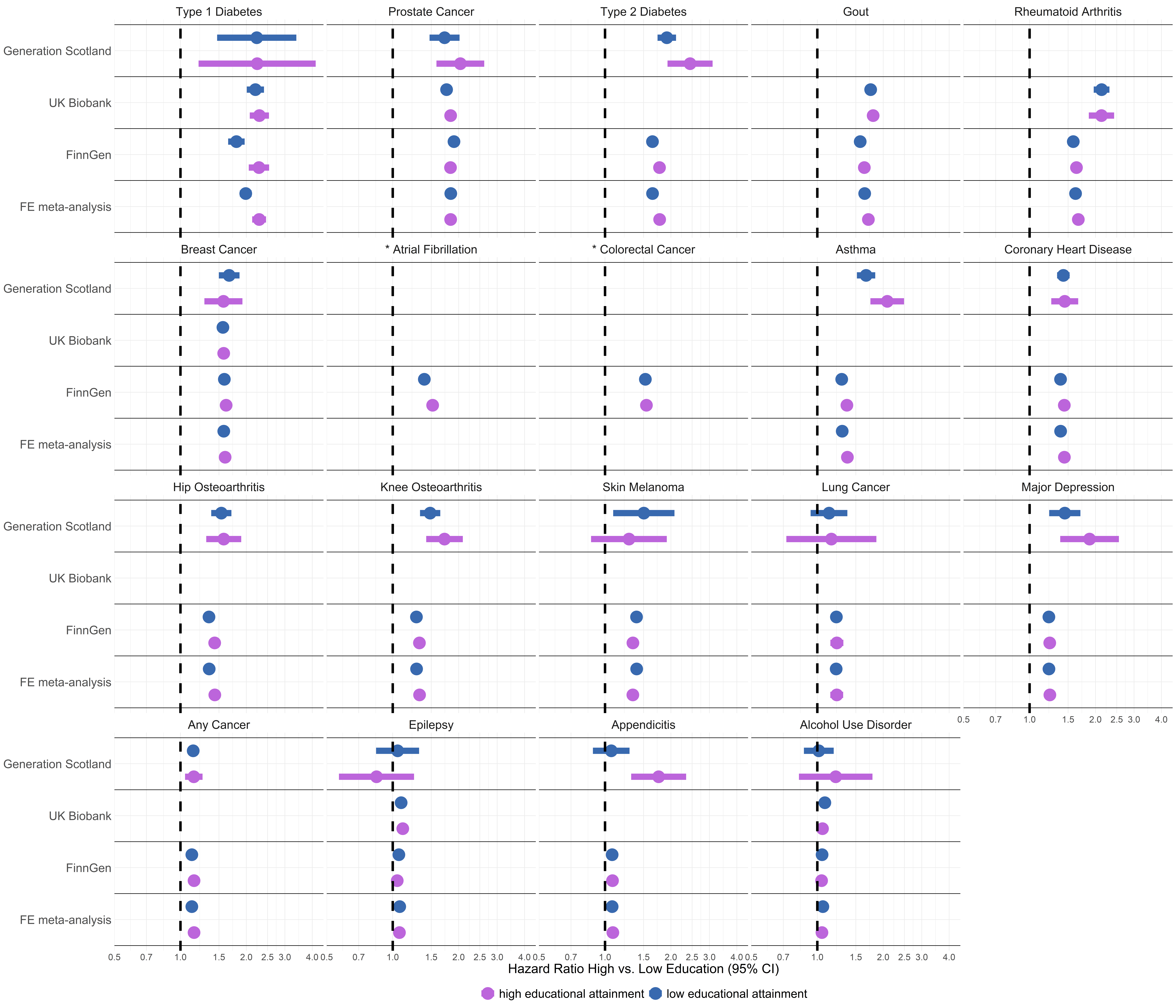

**Supplementary Fig. 7.** Educational attainment-specific cumulative incidence estimates in FinnGen. Bootstrapped 95% confidence intervals reflect the uncertainty of the cumulative incidence estimates for the top, median, and bottom of the disease-specific PGS distribution for (**A**) asthma, (**B**) any cancer, (**C**) coronary heart disease, (**D**) hip osteoarthritis, (**E**) knee osteoarthritis, (**F**) prostate cancer, and (**G**) type 1 diabetes. The exact values are in **Supplementary Table 12**, the exact values of the underlying Cox proportional hazard model are in **Supplementary Table 11**, and the sample sizes by case-control status and educational attainment are in **Supplementary Table 10**.

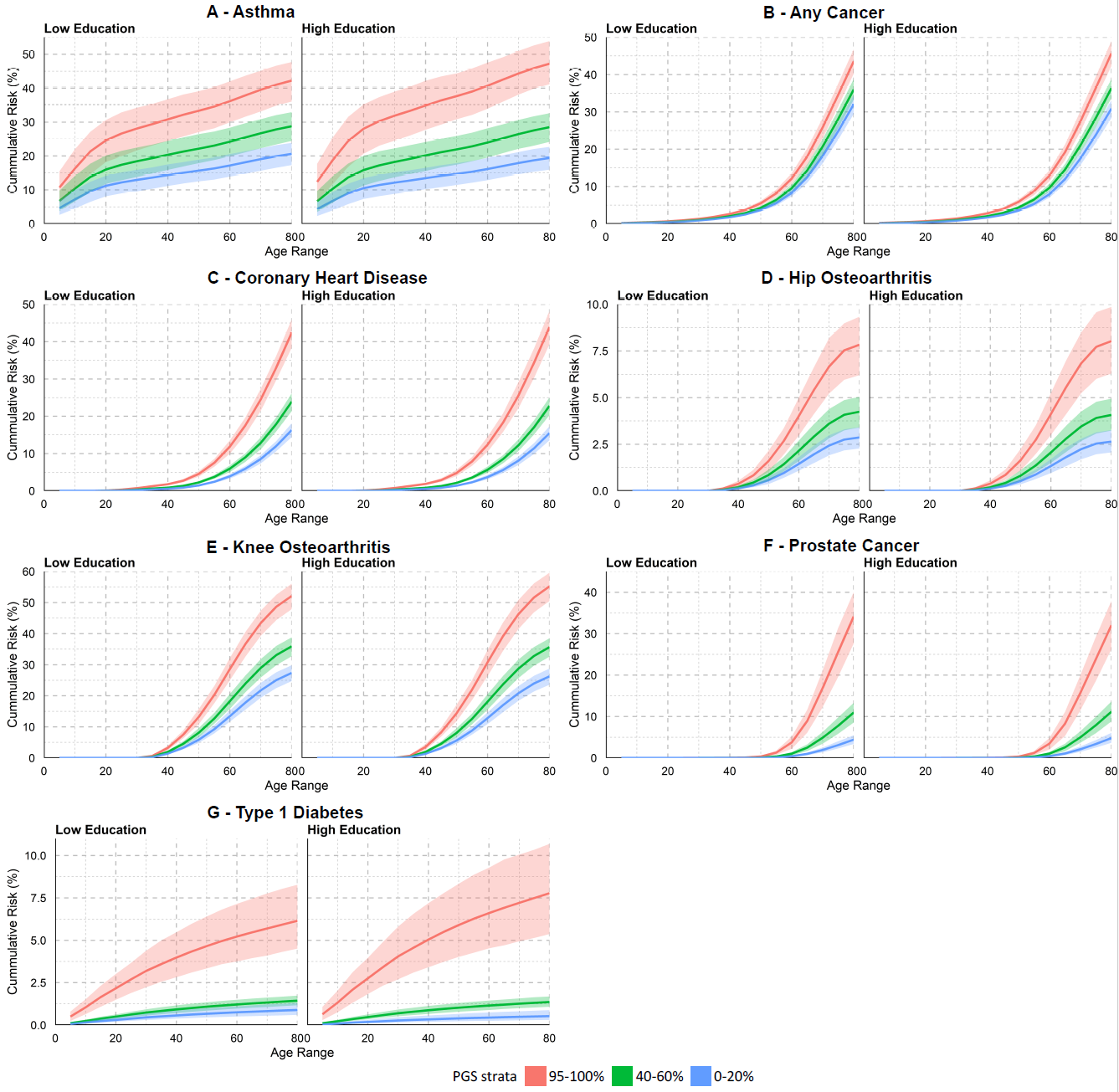

**Supplementary Fig. 8.** Comparison of the hazard ratios per standard deviation of the disease-specific polygenic scores, for individuals with high education relative to individuals with low education, and for the interaction between the disease-specific polygenic scores and educational attainment from Cox proportional hazard (Cox-PH) regression versus Fine-Gray (FG) competing risk models in FinnGen. **A.** Comparison of the hazard ratio for individuals with high education relative to low education in the Cox-PH models (x-axis) versus the FG models (y-axis). **B.** Comparison of the hazard ratios per standard deviation of the disease-specific polygenic scores on disease risk in the Cox-PH models (x-axis) versus the FG models (y-axis). **C.** Comparison of the hazard ratio of the interaction between the disease-specific polygenic score and educational attainment in the Cox-PH models (x-axis) versus the FG models (y-axis). The exact values are in **Supplementary Tables 9** and **22-23**. Sample sizes by case-control status and educational attainment for each complex disease are in **Supplementary Tables 4** and **21**.

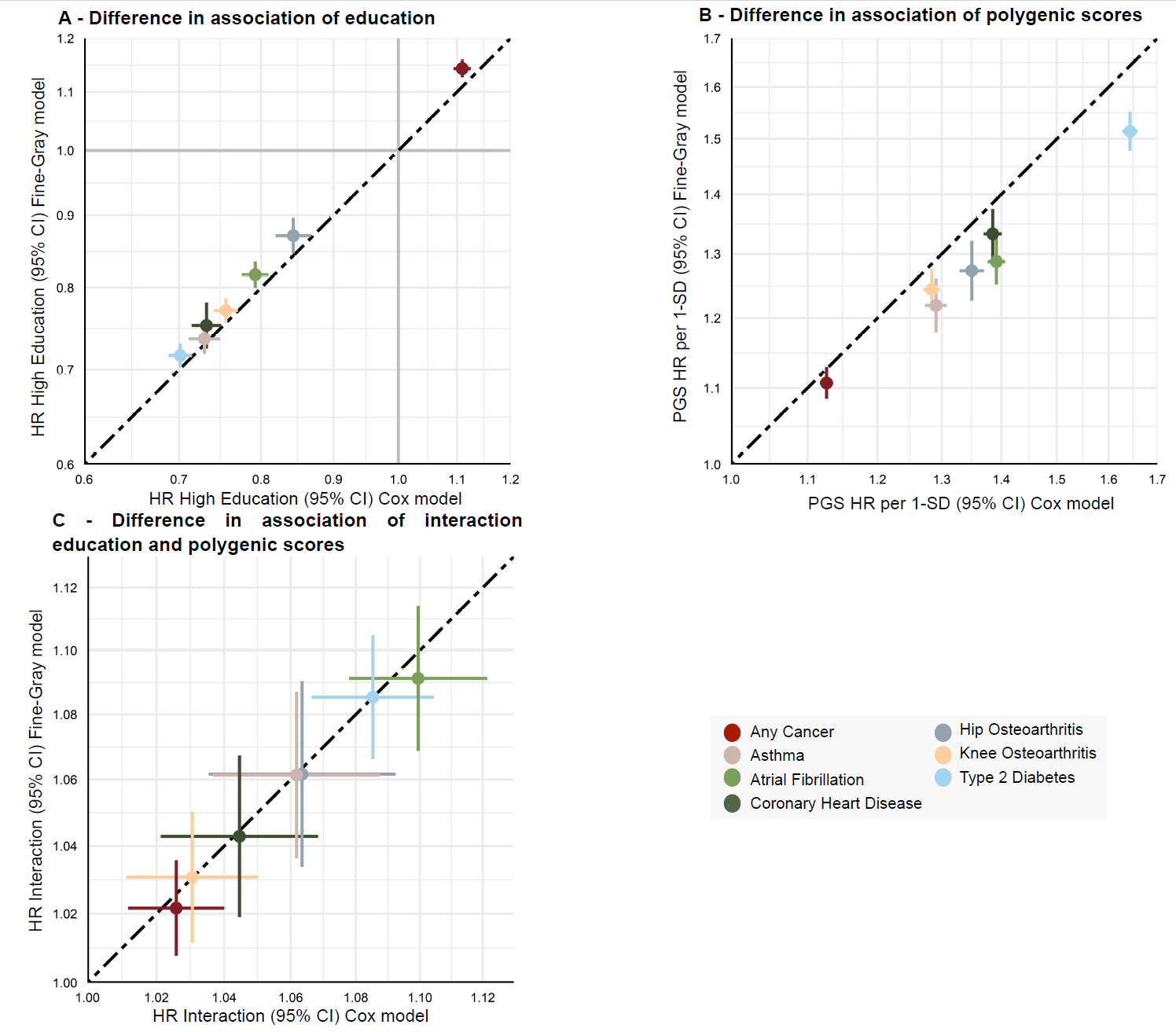

**Supplementary Fig. 9.** Ancestry-specific hazard ratios per standard deviation of the disease-specific polygenic scores (black) and for individuals with high education relative to individuals with low education (purple) on complex disease risk where the disease-specific polygenic scores and education were modelled separately in the UK Biobank. The exact values are in **Supplementary Tables 5** and **25**. Sample sizes by case-control status and educational attainment for each complex disease in are in **Supplementary Tables 4** and **24**.

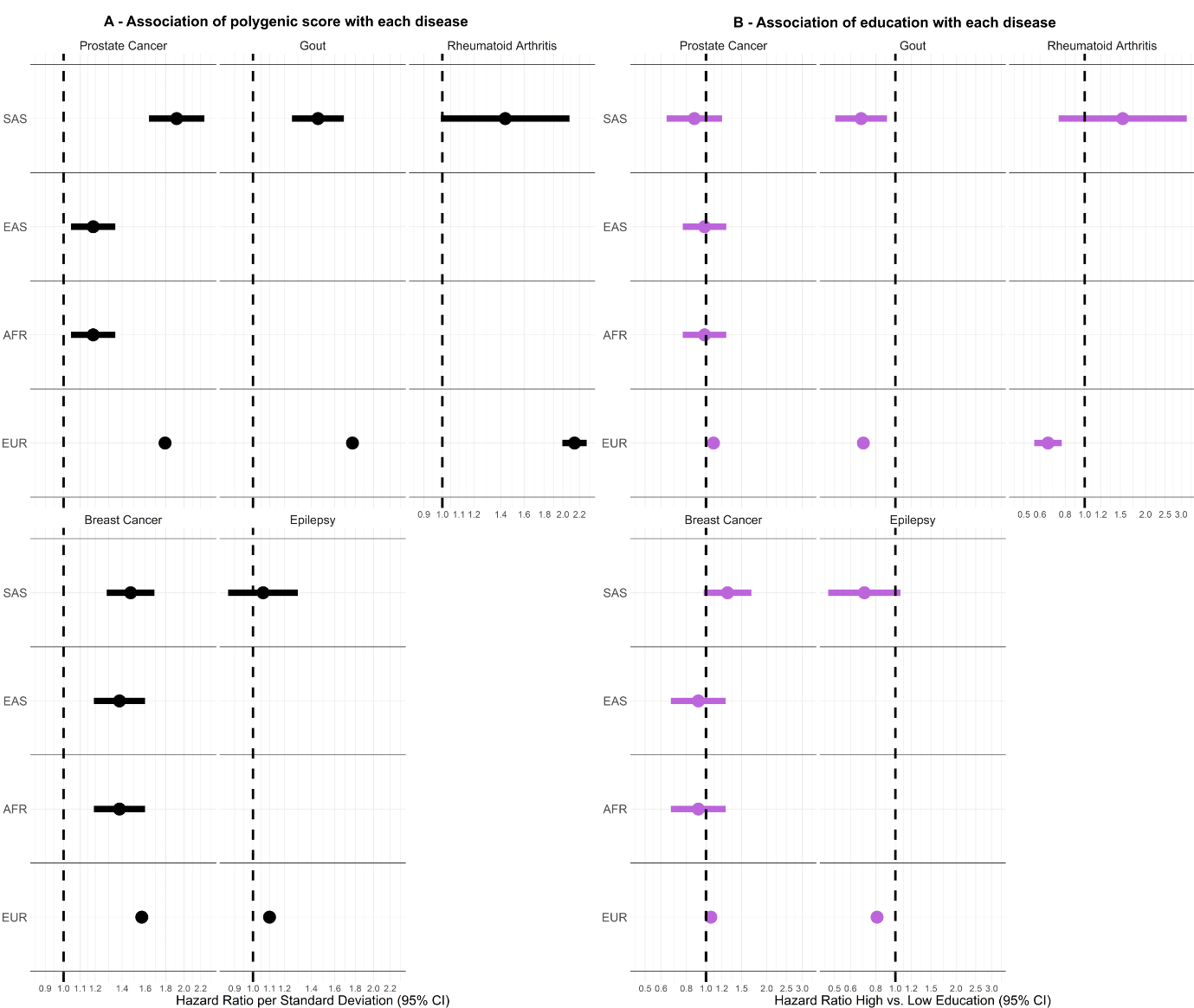

**Supplementary Fig. 10.** Hazard ratios for the relative risk of disease-specific polygenic scores and occupation on 19 complex diseases in FinnGen. **A.** Hazard ratios per standard deviation of the disease-specific polygenic scores on risk of complex diseases. **B**. Comparison of the hazard ratio per standard deviation of the disease-specific polygenic scores on complex disease risk unadjusted (x-axis) and adjusted (y-axis) for the effect of upper-lvel occupation. **C.** Hazard ratios for individuals with upper-level occupation relative to individuals with lower-level occupation on risk of complex diseases. **D**. Comparison of the hazard ratio for individuals with upper-level occupation relative to individuals with lower-level occupation on complex disease risk unadjusted (x-axis) and adjusted (y-axis) for the disease-specific polygenic score**.** The exact values are in **Supplementary Tables 27-29**. Sample sizes by case-control status and occupation for each complex disease are in **Supplementary Table 26**.

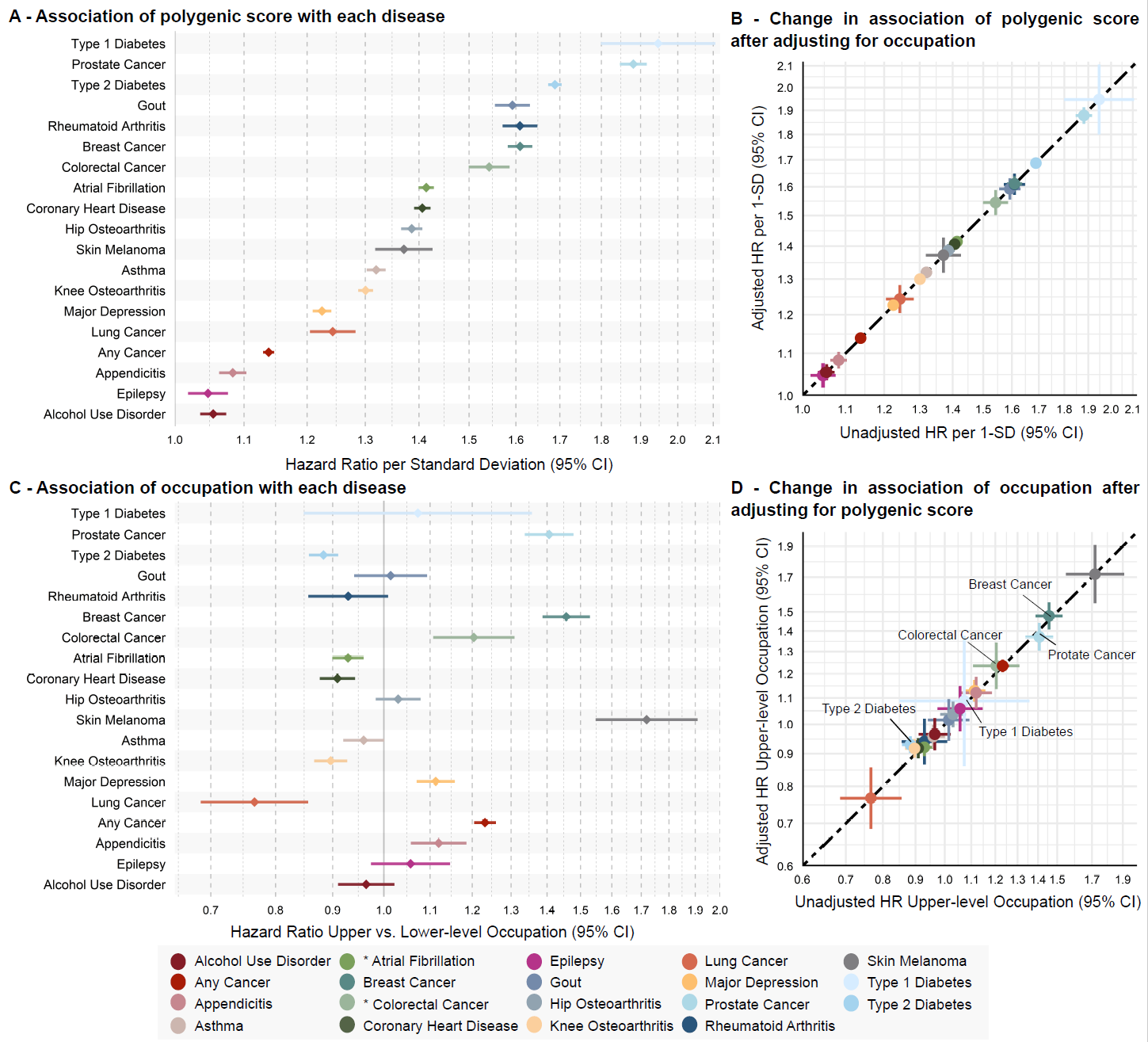

**Supplementary Fig. 11.** Hazard ratios for the relative risk of the disease-specific polygenic scores by educational attainment or occupation on 19 complex diseases in FinnGen. **A.** Hazard ratios per standard deviation of the disease-specific polygenic score in the low (blue) or high (pink) education group. **B.** Hazard ratios per standard deviation of the disease-specific polygenic score in the lower-level (blue) or upper-level (pink) occupation group. Significance of the differences in the effect of the disease-specific polygenic score per education or occupation group evaluated by the statistical significance of the interaction term between the disease-specific PGS and education or occupation level after Bonferroni correction for multiple testing of 19 outcomes (*p* < 2.63x10^-03^). The exact values are in **Supplementary Tables 8-9** and **30-31**. Sample sizes by case-control status and educational attainment or occupation for each complex disease are in **Supplementary Tables 4** and **26.**

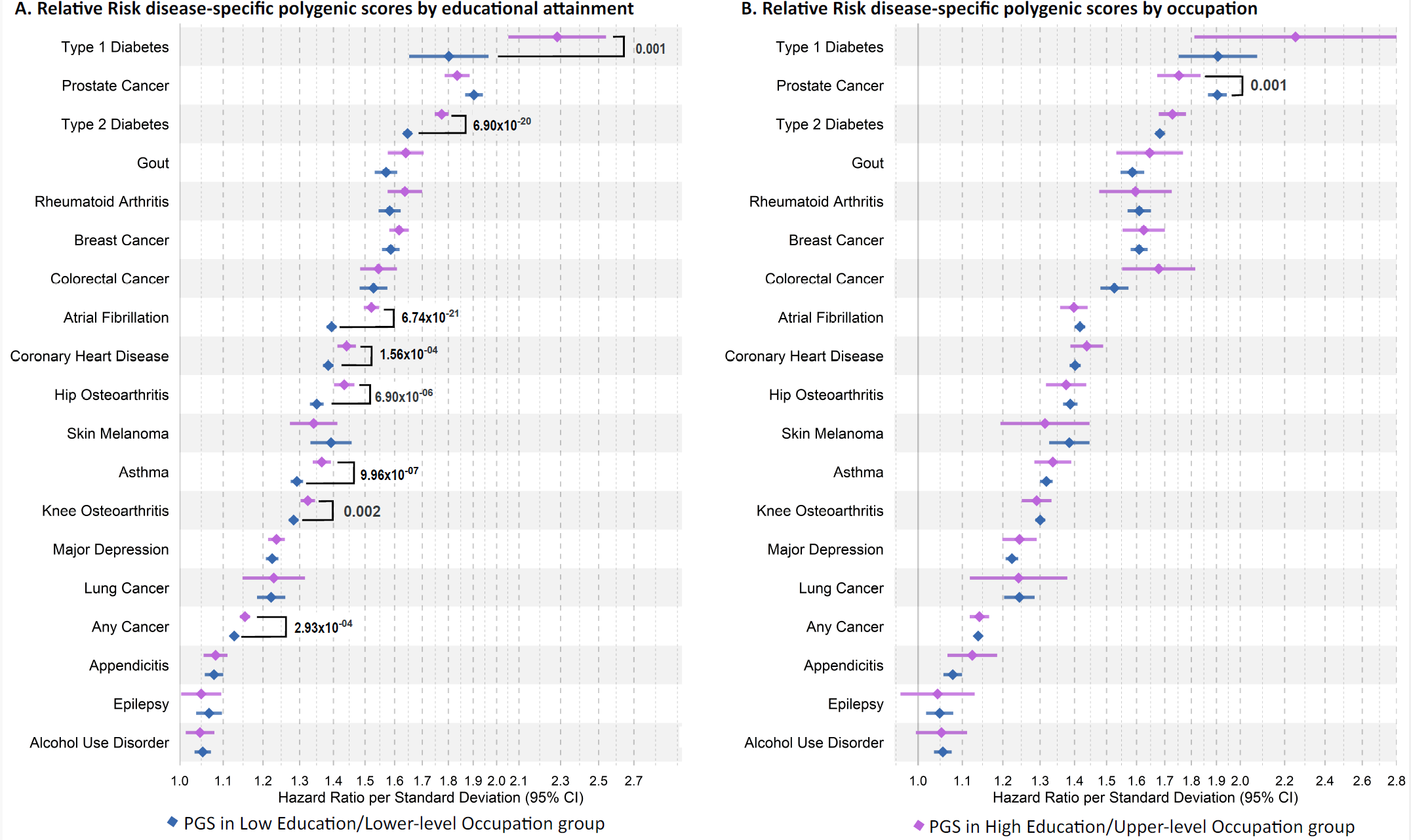

**Supplementary Fig. 12.** Comparison of the hazard ratios across the educational attainment and occupation models in FinnGen. **A.** Comparison of the hazard ratio for individuals with high education relative to individuals with low education on complex disease risk (x-axis) against hazard ratio for individuals with upper-level occupation relative to individuals with lower-level occupation on complex disease risk (y-axis). **B.** Hazard ratio per standard deviation of the disease-specific polygenic scores on complex disease risk in individuals with educational attainment available (x-axis) compared to individuals with occupation available (y-axis). **C**. Comparison of the hazard ratio per standard deviation of the disease-specific polygenic score in the low education (x-axis) versus lower-level occupation (y-axis) groups. **D.** Comparison of the hazard ratio per standard deviation of the disease-specific polygenic score for individuals in the high education (x-axis) versus upper-level occupation (y-axis) group. The exact values are in **Supplementary Tables 5-6**, **8**, **27-28**, and **30**. Sample sizes by case-control status and educational attainment or occupation for each complex disease are in **Supplementary Tables 4** and **26**.

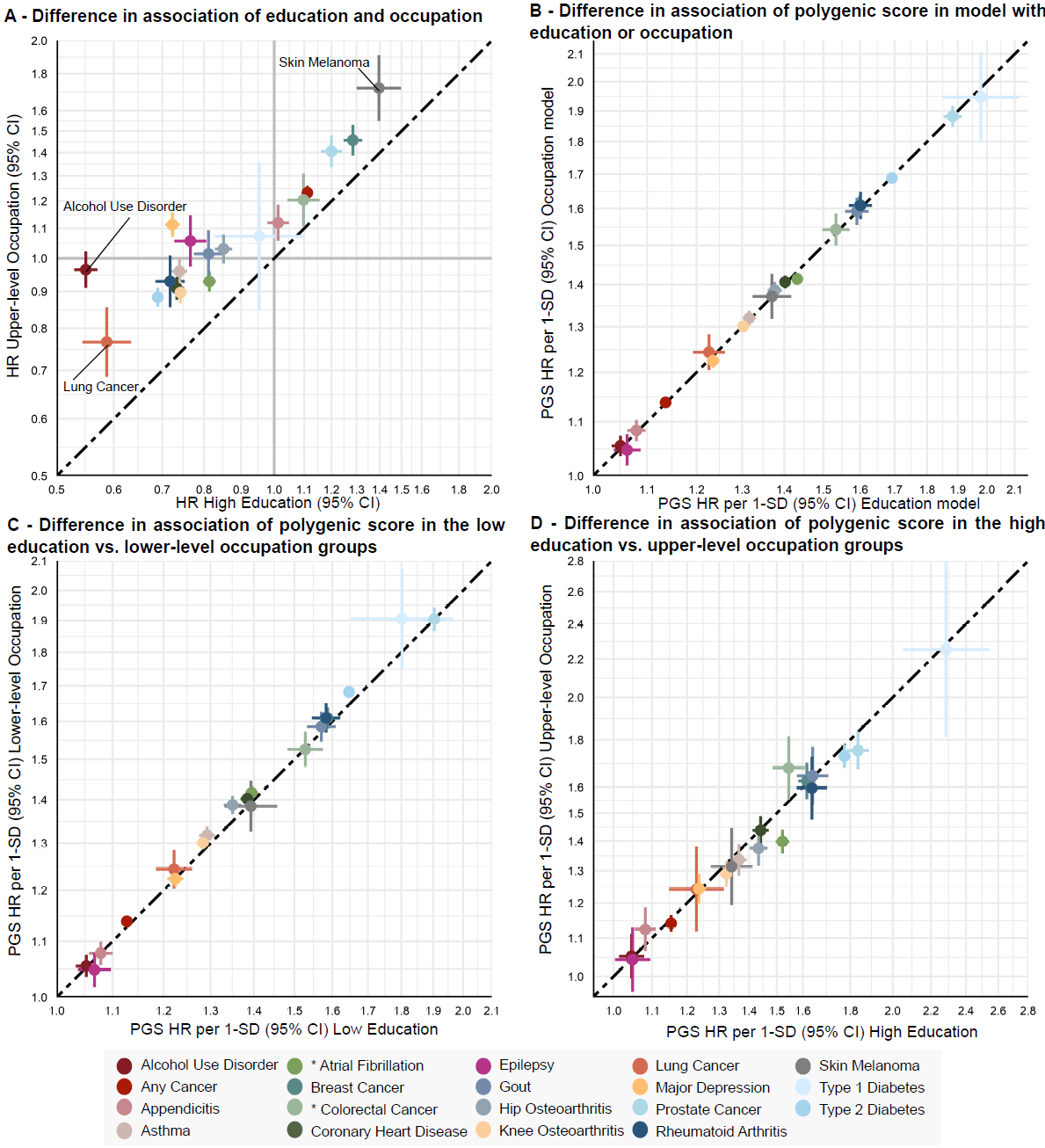

**Supplementary Fig. 13.** Occupation-specific cumulative incidence estimates in FinnGen. Bootstrapped 95% confidence intervals reflect the uncertainty of the cumulative incidence estimates for the top, median, and bottom of the disease-specific PGS distribution for (**A**) asthma, (**B**) any cancer, (**C**) coronary heart disease, (**D**) hip osteoarthritis, (**E**) knee osteoarthritis, (**F**) prostate cancer, (**G**) type 2 diabetes, and (**H**) atrial fibrillation. The exact values are in **Supplementary Table 34**, the exact values of the underlying Cox proportional hazard model are in **Supplementary Table 33**, and the sample sizes by case-control status and occupation are in **Supplementary Table 32**.

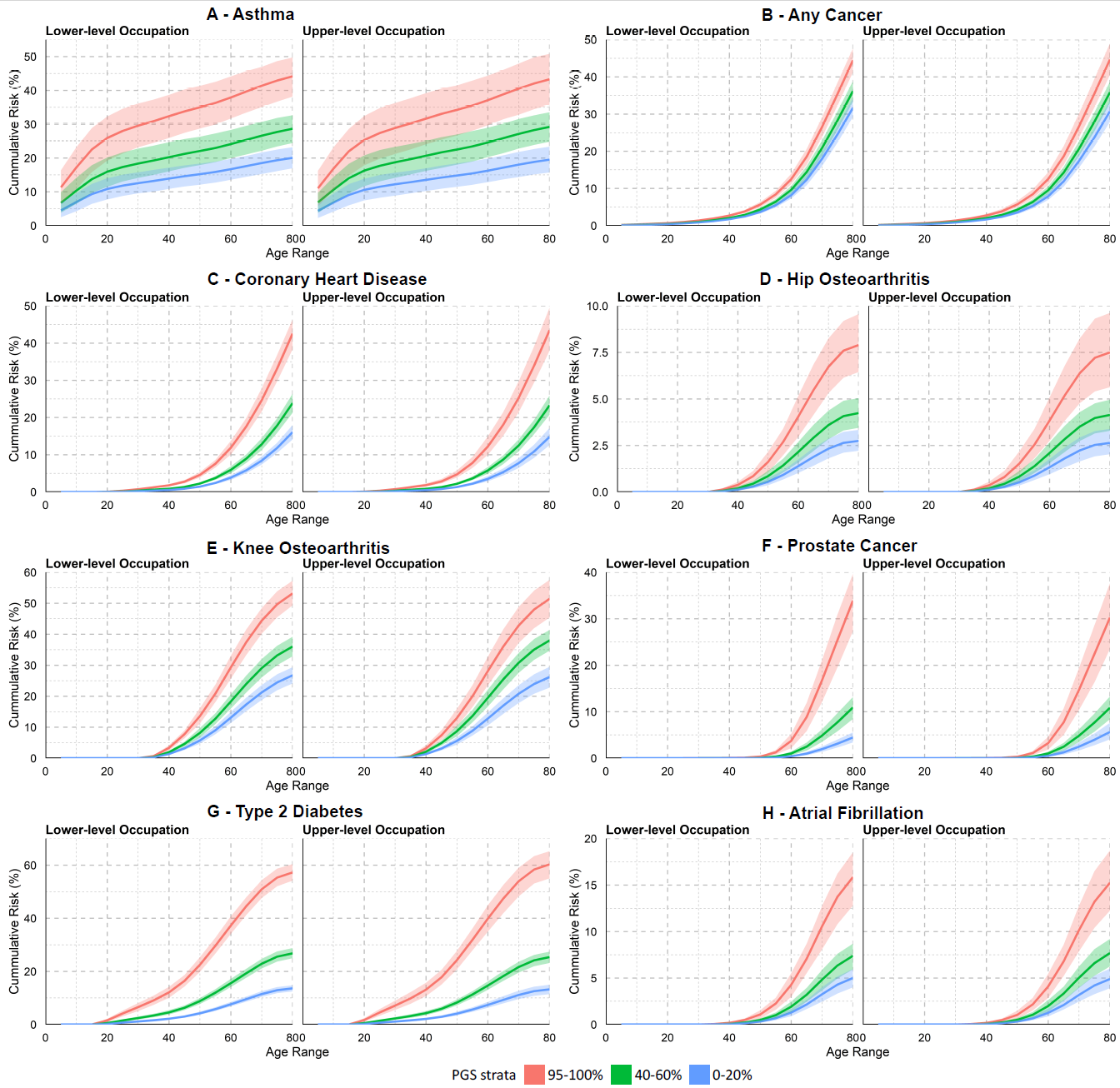

### **Supplementary References**

1.	Kurki, M. I. *et al.* FinnGen provides genetic insights from a well-phenotyped isolated population. *Nature* **613**, 508–518 (2023).

2.	Zhao, S. *et al.* Strategies for processing and quality control of Illumina genotyping arrays. *Brief Bioinform* **19**, 765–775 (2017).

3.	Chang, C. C. *et al.* Second-generation PLINK: rising to the challenge of larger and richer datasets. *GigaScience* **4**, s13742-015-0047–8 (2015).

4.	Loh, P.-R. *et al.* Reference-based phasing using the Haplotype Reference Consortium panel. *Nat Genet* **48**, 1443–1448 (2016).

5.	Sequencing Initiative Suomi project (SISu), Institute for Molecular Medicine Finland (FIMM), University of Helsinki, Finland. *SISU v4.1* https://sisuproject.fi/.

6.	Auton, A. *et al.* A global reference for human genetic variation. *Nature* **526**, 68–74 (2015).

7.	Bellenguez, C. *et al.* A robust clustering algorithm for identifying problematic samples in genome-wide association studies. *Bioinformatics* **28**, 134–135 (2012).

8.	Bycroft, C. *et al.* The UK Biobank resource with deep phenotyping and genomic data. *Nature* **562**, 203–209 (2018).

9.	Galinsky, K. J. *et al.* Fast Principal-Component Analysis Reveals Convergent Evolution of *ADH1B* in Europe and East Asia. *The American Journal of Human Genetics* **98**, 456–472 (2016).

10.	Howie, B., Fuchsberger, C., Stephens, M., Marchini, J. & Abecasis, G. R. Fast and accurate genotype imputation in genome-wide association studies through pre-phasing. *Nat Genet* **44**, 955–959 (2012).

11.	McCarthy, S. *et al.* A reference panel of 64,976 haplotypes for genotype imputation. *Nat Genet* **48**, 1279–1283 (2016).

12.	Huang, J. *et al.* Improved imputation of low-frequency and rare variants using the UK10K haplotype reference panel. *Nat Commun* **6**, 8111 (2015).

13.	Pain, O., Al-Chalabi, A. & Lewis, C. M. The GenoPred pipeline: a comprehensive and scalable pipeline for polygenic scoring. *Bioinformatics* **40**, btae551 (2024).

14.	Gibbs, R. A. *et al.* The International HapMap Project. *Nature* **426**, 789–796 (2003).

15.	Price, A. L. *et al.* Long-Range LD Can Confound Genome Scans in Admixed Populations. *The American Journal of Human Genetics* **83**, 132–135 (2008).

16.	Durbin, R. Efficient haplotype matching and storage using the positional Burrows–Wheeler transform (PBWT). *Bioinformatics* **30**, 1266–1272 (2014).

17.	Delaneau, O., Zagury, J.-F. & Marchini, J. Improved whole-chromosome phasing for disease and population genetic studies. *Nat Methods* **10**, 5–6 (2013).

18.	Delaneau, O., Marchini, J. & Zagury, J.-F. A linear complexity phasing method for thousands of genomes. *Nat Methods* **9**, 179–181 (2012).

19.	O’Connell, J. *et al.* A General Approach for Haplotype Phasing across the Full Spectrum of Relatedness. *PLOS Genetics* **10**, e1004234 (2014).

20.	Gray, A., Stewart, I. & Tenesa, A. Advanced Complex Trait Analysis. *Bioinformatics* **28**, 3134–3136 (2012).
